# Nonlinear dynamic genetic regulation identifies peripheral drivers of neurodegenerative disease progression

**DOI:** 10.64898/2026.01.29.26345161

**Authors:** Junfeng Luo, Hao Wu, Wenxuan Du, the Alzheimer’s Disease Neuroimaging Initiative, Ganqiang Liu

**Author notes:** Correspondence should be addressed to: Ganqiang Liu, Ph.D., School of Medicine, Sun Yat-sen University, 66 Gongchang Road, Guangming District, Shenzhen, Guangdong, 518107, China. Data used in preparation of this article were obtained from the Alzheimer’s Disease Neuroimaging Initiative (ADNI) database (adni.loni.usc.edu). As such, the investigators within the ADNI contributed to the design and implementation of ADNI and/or provided data but did not participate in analysis or writing of this report. A complete listing of ADNI investigators can be found at: http://adni.loni.usc.edu/wpcontent/uploads/how_to_apply/ADNI_Acknowledgement_List.pdf.

## Abstract

The dynamic genetic regulation of gene expression during the progression of common neurodegenerative diseases (NDDs) remains poorly characterized, obscuring the genetic architecture of longitudinal clinical traits. Here, we present 2sGen-GPS, a two-stage genetic Granger temporal causality framework designed to integrate genetic variants, intermediate longitudinal molecular traits, and disease progression phenotypes. In the discovery stage, we utilized multivariate polynomial temporal genetic association (MPTGA) analysis of peripheral blood to identify 774,533 time-dependent cis-eQTLs (11,936 eGenes), enabling the imputation of individual-level longitudinal expression trajectories. In the second stage, these imputed profiles were linked to NDD phenotypes to infer temporal causal relationships. Applying 2sGen-GPS to Parkinson’s disease multi-omics cohorts, we identified a peripheral-to-central regulatory axis; specifically, the rs11241912-driven temporal expression of *C5orf63* exhibits a lagged causal association with cerebrospinal fluid LRP1 levels and motor symptom progression. Cross-regional single-cell analysis further revealed that rs11241912 carriers harbor a localized compensatory signature in excitatory neurons of the globus pallidus interna, characterized by the up-regulation of dopamine receptor signaling pathways. Extending 2sGen-GPS to Alzheimer’s disease, we identified 328 genes linked to cognitive decline and prioritized drug compounds capable of reversing these expression signatures. Our study elucidates how dynamic genetic effects shape the trajectories of NDD-related traits and nominates peripherally accessible, causal genes as promising therapeutic targets for modulating disease progression.

## Introduction

Common aging-associated neurodegenerative diseases (NDDs), such as sporadic Parkinson’s disease (PD) and late-onset Alzheimer’s disease (AD), are defined by their protracted clinical courses and profound phenotypic heterogeneity^1,2^. Certain biological processes, such as immune responses, are dynamic and often most prominent in the early stages of disease. Therefore, elucidating the genetic architecture of longitudinal complex traits in NDDs is crucial for clarifying these temporal biological changes and optimizing the timing of clinical trials targeting specific pathways^3^. While large-scale genome-wide association studies (GWAS) have identified numerous risk loci for PD^4^ or AD^5^, and expression quantitative trait locus (eQTL) analyses have helped link genotypes to molecular phenotypes, the vast majority of these efforts rely on cross-sectional “snapshot” data^6,7^. Such static approaches fail to capture the evolving nature of genetic regulatory effects across the disease continuum, leaving the temporal genetic basis of PD and AD progression largely unresolved.

Current strategies for integrating eQTL and GWAS, including Mendelian randomization (MR)^8-11^, co-localization^9,12^, TWAS^13^ or UTMOST^7^, have successfully bridged genetic variants with intermediate traits in static contexts. However, analytical frameworks capable of deconstructing temporal genetic regulation and linking it to longitudinal trajectories remain a critical unmet need. To address this gap, we developed 2sGen-GPS, a two-stage genetic Granger temporal causality framework. By employing a multivariate polynomial temporal genetic association (MPTGA) approach, we identified nonlinear, dynamic eQTLs within longitudinal PD cohorts that eluded detection in previous cross-sectional analyses^14,15^. Our framework identified key dynamic driver genes - including a novel peripheral-to-central regulatory axis involving the rs11241912 variant - that modulates symptom progression via CSF mediators and cell-type-specific responses in the globus pallidus interna (GPI). These findings offer a high-resolution map of the dynamic genetic mechanisms underlying neurodegeneration and nominate time-sensitive therapeutic targets for halting disease progression.

## Methods

### Datasets

We collected comprehensive datasets comprising whole genome sequencing (WGS) genotypes (*n*=3,565), longitudinal RNA-seq samples (*n*=8,253), longitudinal targeted proteomics datasets (*n*=3,050) and longitudinal phenotypes from the Accelerating Medicines Partnership in Parkinson’s Disease (AMP-PD) consortium (including PPMI^16^, PDBP^17^ and SURE-PD3 cohorts, *https://amp-pd.org*). Additionally, longitudinal T1w MRI brain images (*n*=1,848) were obtained from the PPMI database (*www.ppmi-info.org*). Furthermore, WGS genotypes (*n*=808) and longitudinal phenotypes data used in preparation of this article were obtained from the ADNI database (*adni*.*loni*.*usc*.*edu*). The ADNI was launched in 2003 as a public-private partnership, led by Principal Investigator Michael W. Weiner, MD. The primary goal of ADNI has been to test whether serial magnetic resonance imaging (MRI), positron emission tomography (PET), other biological markers, and clinical and neuropsychological assessment can be combined to measure the progression of mild cognitive impairment (MCI) and early Alzheimer’s disease (AD). For up-to-date information, see www.adni-info.org. Unimputed genotypes (*n*=1,709) for the ROSMAP^18^ cohort, utilizing the Affymetrix GeneChip 6.0, were downloaded from the Synapse website (id: syn17008939), while the longitudinal phenotypes for the MAP cohort in ROSMAP were accessed from the RADC Research Resource Sharing Hub (*https://www.radc.rush.edu*). See the website of these cohorts for details regarding the inclusion and exclusion criteria of participants.

All cohorts included in this study enrolled participants with informed consent and collected and analyzed data with approval from the local ethics committees. The Institutional Review Board of the School of Medicine, Sun Yat-sen University approved the current analyses.

### Dataset partitioning and data preprocessing

We selected a total of 639 healthy controls and PD cases from the AMP-PD with genotype data and five follow-up visits with peripheral blood transcriptomics data (Total RNA sequencing). Within this dataset, 293 participants from the PPMI were assigned as the discovery dataset, contributing to a total of 1,465 longitudinal visits over a three-year period (Figure S2a-b). The remaining 346 participants from the PDBP were assigned as the independent replication dataset, accounting for a total of 1,730 longitudinal visits spanning two years (Figure S2c-d). Each participants contained five longitudinal time points.

The WGS data was generated using the Illumina HiSeq XTen squencer with samples coming from whole blood. Primary preprocessing and quality control procedures for the genotype data were demonstrated in the AMP-PD website (*https://amp-pd.org/whole-genome-data*). We applied quality control (QC) for subjects (genotype success rate > 95%, genotype-derived gender concordant with reported gender) and for single nucleotide polymorphisms (SNPs) (Hardy-Weinberg equilibrium p < 1×10^-6^); minor allele frequency (MAF) > 0.05, genotype call rate > 0.95) on each cohort separately. We computed genotype principal components (PCs) based on the post-sample and variant QC WGS. QC and genotype PCs computed was performed using *PLINK 1*.*9*. We utilized the initial eight genotype PCs that were statistically significant (Table S1, S2) to account for population stratification on each dataset. We employed *EIGENSTRAT* (v6.1.4) to calculate the number of statistically significant PCs^19^.

Total RNA-seq samples from both cohorts were extracted from whole blood. Details of transcriptome sequencing (LongRNA-seq) procedure, alignment, initial quality control and quantification were also listed at AMP-PD website (*https://amp-pd.org/transcriptomics-data*). We downloaded the longitudinal RNA gene expression data from AMP-PD. Transcripts per million (TPM) was used to measure expression level. In the discovery cohort, transcripts with a width of transcripts less than 200nt and a transcript level less than 1 mean value were removed. In total of 17,236 autosomal gene-expression traits were set aside and log2 transformed for further analysis.

To reduce the occurrence of the spurious correlation in subsequent temporal association analysis, we test the stationarity of longitudinal series data on the 17,236 gene-expression traits from multiple individuals based on the Augmented Dickey-Fuller (ADF) unit root test^20^ (Supplemental Methods). *P*<0.05 were considered as significantly indicating that the time series is stationary. In total, 17,222 stationary longitudinal gene-expression traits passed the ADF test with the *P* < 0.05.

To accounted for hidden technical confounders (such as batch effects) and biological confounders (such as inter-individual differences in cell type composition or transcriptome-wide variance in the gene expression data), we used the first 33 transcriptome-wide expression principal components (PCs) on the discovery cohort and 37 PCs on the replication cohort as the latent covariates for gene expression levels (Figure S4a,c, Supplemental Methods).

### MPTGA, method for temporal eQTL mapping

Stationary temporal gene-expression trait of subject *i* were modelled using cubic polynomial function with respect to a given genetic locus, in terms of a covariates adjusted model as

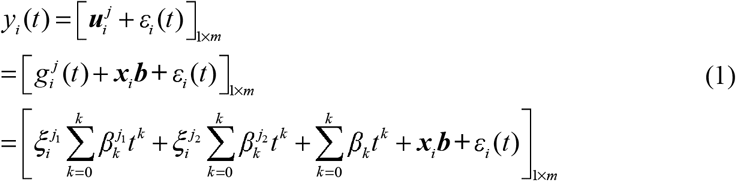

Where 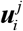 is the mean vector for genotype *j* of individual *i* with multivariate distribution and *m* is the number of time points in time-series data; 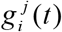 is the temporal genetic component including genetic and interaction between genetic and time terms for genotype *j* of individual *i* ; ***x***_*i*_ is a (m×n)-design matrix of individual *I* for *n* covariates, ***b*** is the vector for *n* covariate effects. Binary encoded genotype 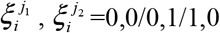 with more sensitive of dynamic eQTL identification (Supplemental Methods) is used for each SNP. Follow-up time ***t*** = [*t*_1_, *t*_2_,K, *t*_*m*_ ], *k* is the order of *t* and *K* = 3 for cubic polynomial nonlinear regression model.

Similar to Ma et al.^21^ and Lin et al.^22^, we assumed that the longitudinal trait followed a multivariate normal density as

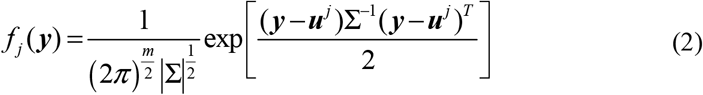

The variance at each time point exists correlation stationarity as

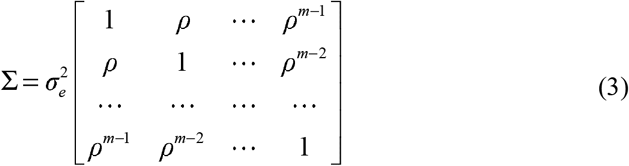

Where ***ρ*** is auto-regression proportion which ranges from 0 to 1. The likelihood for temporal trait can be written as

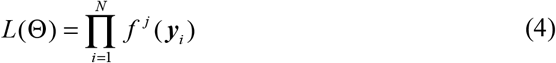

Where 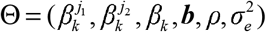 is the set of unknown parameters in the statistical model. Maximum likelihood estimates (MLEs) were calculated by taking derivative of log *L* (Θ ) with respect to each unknown parameter. See Supplemental Methods about the detail of parameters estimation.

After determining parameters with MLE procedure, the significance of temporal eQTL, including total effects of genotype and interaction between genotype and time on trait *y*, can be tested by comparing a null model (single temporal trait curve) *H*_*0*_:

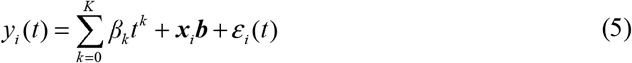

vs. a full model (different temporal trait curve for different genotypes) *H*_*1*_:

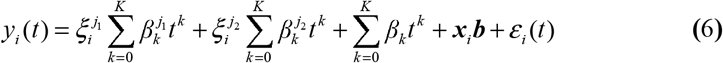

to access whether the existence of temporal eQTL.

The test statistic for testing the above hypotheses is calculated as the log-likelihood ratio (LR) of the full model (*H*_*1*_) over the reduced model (*H*_*0*_)

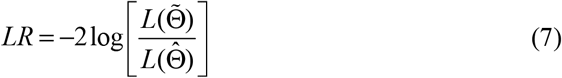

Where 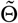 % and 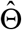 denote the ML estimates of the unknown parameters under *H*_*0*_ and *H*_*1*_, respectively. The test statistics for the LR test can be viewed as being asymptotically *χ*^2^ distributed with 8 d.f.

### Method of dynamic eQTL mapping

Dynamic eQTL, whose effect varies significantly over time, can be detected by testing the significance of interaction effect between genotype and time by comparing a null model (without interaction between QTL and time) *H*_*0*_

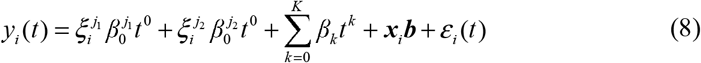

vs. full model as equation (6).

The test statistic for testing the above hypotheses is also conducted based on LR test. The test statistics for the LR test here can be viewed as being asymptotically *χ*^2^ distributed with 6 d.f. An eQTL without significant interaction *P* value with time is assigned as additive eQTL.

### Proportion of variance in phenotype explained (PVE) by temporal eQTL

The computation of PVE^23^ can be presented as

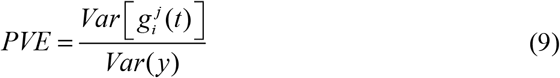

Where *Var(y)* is the variance of phenotype y, 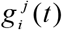 is temporal eQTL mean vector in equation (1).

### Power estimate

We conducted bootstrap to generate the distribution of LR (Alternate hypothesis, H1, distribution. Figure S6a) in equation (7) to estimate the type-II error, *β*, of a temporal eQTL under a specific statistical significance cutoff, *α*. Then the power of temporal eQTL can be expressed as power = 1-*β*. The estimate for LR of each temporal eQTL was evaluated using 1,000 bootstraps where subjects’ ID was sampled with replacement and LR was recomputed each time. Power estimate was conducted on 5,000 temporal *cis*-eQTLs randomly selected from the temporal *cis*-eQTLs detected in MPTGA and samples sizes.

### Local temporal eQTL mapping

We initially performed temporal *cis*-eQTL mapping on the 17,222 stationary longitudinal gene-expression traits in the discovery dataset, adjusted by age at baseline, male sex, disease diagnosis, RNA integrity number (RIN) and eight genotype PCs (Table S1, S2) that were not fully explained by the gene expression PCs (Figure S4b, d). Genes with a window 1 Mb upstream and 1 Mb downstream around each SNP were selected for testing. This resulted in a total of 77,631,179 SNP-gene pairs for temporal *cis*-eQTL mapping. We computed Benjamini and Hochberg FDR^24^ to account for multiple testing at all gene-SNP pairs. SNP-gene pairs with *FDR* < 0.05 (raw *P* = 0.001) were considered as temporal eQTL and subsequently validated in an independent dataset to confirm these findings (*P*<0.05). Additionally, we calculated the PVE by a given SNP for all temporal *cis*-eQTLs found in both the discovery and replication datasets.

### Local nonlinear dynamic eQTL mapping

Subsequently, nonlinear dynamic *cis*-eQTL mapping were performed on the temporal *cis*-eQTLs, and significance was determined by *FDR* < 0.05 in the discovery dataset and *P*<0.05 in the replication dataset. The temporal *cis*-eQTLs with *P* > 0.05 in both datasets were categorized as additive *cis*-eQTL.

### Comparison between MPTGA and existing methods for temporal eQTL mapping

We compared MPTGA and existing temporal eQTL mapping approaches for local temporal eQTL mapping in the simulated and real-world data (Supplemental Methods). Temporal *cis*-eQTL mapping in the Union was performed using *FastQTL* (v2.0), while the other three approaches were performed using the functions in our newly developed tool *2sGen-GPS*. Age at baseline, male sex, diagnosis, RIN value, 8 population structure PCs and 33 expression PCs were considered as covariates for all methods. A significant *P* value cutoff was determined by an FDR < 0.05 to identify genes with significant temporal eQTL. Linear or nonlinear dynamic *cis*-eQTL mapping was also carried out for the LR and Cubic approaches.

### Two-stage genetic Granger temporal causality study

We established a two-stage genetic Granger temporal causality study (2sGen-GPS) framework to determine whether a longitudinal gene-phenotype association exhibits temporal causality. This framework combines the idea of two-stage least squares (2SLS) MR^8,25^ and the Grander causality test^26,27^. The Granger causality test aims to test whether the variance of the prediction of a temporal phenotype *Y* could be significantly reduced by incorporating information from the history of another time series *X*, and thus to calculate the lagged correlation between *X* and *Y* to test whether *X* is deemed to be “causal” of *Y*. Furthermore, the use of instrumental variable (IV) approaches has been employed to strengthen causal inferences in non-experimental situations^8^.

In 2sGen-GPS, for a given gene with at least one temporal *cis*-eQTL, its imputed temporal expression level *E* by a single temporal eQTL SNP IV and the longitudinal phenotype *Y* can be modeled by linear vector autoregressive (VAR) model:

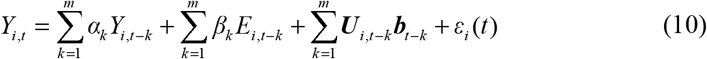

to explore the Granger temporal causal relationship between two temporal traits.

Where *Y*_*i,t*_ and *Y*_*i,t* −*k*_ are the future and past lagged observations of trait *Y* of individual *i, k* is the order or lag of a lagged time series which maximum lag is *m*; α*k* and β_*k*_ are the effect size of *k* order lagged correlation effect size; *E*_*i,t* −*k*_ is the lagged predicted temporal expression level of gene *E* which is presented as temporal genetic component 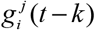 in equation (1); ***U***_*i,t*−*k*_ is a matrix of individual *i* for *n* latent confounds, ***b***_*t* −*k*_ is the vector for *n* confound effects; *ε*_*i*_ (*t* ) is the white noise or error term with *N* (0, *σ* ^2^ ) . We performed an ordinary linear regression to estimate the parameters in equation (10) and used an *F-test* to compare the null model 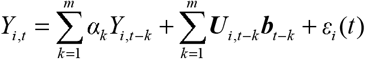 against the full model in equation (10) to detect temporal causal effect. In brief, the 2sGen-GPS framework consists of the following two stages:

1. Individual temporal gene expression imputation by a genetic IV. The genetic IV corresponding to a temporal eQTL gene should formally satisfy the following assumptions^8^: I. The IV *Z* is associated with the exposure of interest *X*; II. *Z* is independent of the confounding factors *U* that confound the association of *X* and the outcome *Y*; III. *Z* is independent of outcome *Y* given *X* and the confounding factors *U*. The parameters in 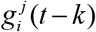 in equation (1) of genetic IV predictor for temporal gene expression imputation are trained in the discovery dataset using the MPTGA method.
2. Estimating temporal causal effect of genetical predicted temporal gene expression level on disease-related longitudinal phenotype.

### Temporal causal effect of genes on PD progression related phenotype

We conducted 2sGen-GPS to investigate potential temporal causal effects of genes with dynamic or additive *cis*-eQTLs on the longitudinal MDS-UPDRS I-III^28,29^ scores in PD following the workflow in Figure S9 . The MDS-UPDRS I-III^28,29^ scores are commonly employed for the clinical evaluation of motor and non-motor symptoms in the PD.

A total of 745 PD cases with 7,478 visits, comprising genotype and clinical phenotype data from the PPMI cohort were included as the discovery dataset (Figure S8). Initially, 1,252 dynamic *cis*-eQTL SNPs were selected to predict 639 longitudinal gene expressions. Subsequently, 602 dynamic *cis*-eQTL SNPs associated with confounders were excluded. A total of 650 dynamic *cis*-eQTL SNPs were used as genetic IV predictors for the imputation of 406 longitudinal gene expressions (with 777 dynamic *cis*-eQTLs). The 2sGen-GPS was then employed to identify potential temporal causal effects of the 406 dynamic *cis*-eQTL genes on longitudinal MDS-UPDRS I-III scores in the discovery dataset. Bonferroni correction was applied to adjust for multiple testing within each lag (m =1,2,3,4,5) with significance set at < 0.05/777 = 6.4×10^-5^. Only dynamic *cis*-eQTLs with significant 2sGen-GPS results and passing the cointegration test^30^ (Supplemental Methods) with *P* < 0.05 were retained.

The findings were subsequently replicated in an independent dataset comprising 680 PD cases with 4,057 visits from the PDBP and SURE-PD3 cohorts (Figure S8) to confirm the temporal causal genes. Dynamic *cis*-eQTLs with raw *P* < 0.05 were considered significant. For a dynamic *cis*-eQTL that showed significance in VAR models with different lags, the model with the minimum SC^31,32^ value (Supplemental Methods) in the replication dataset was retained. Additionally, *cis*-eQTLs with a distinct direction of causal effect size of the first-order lagged series term in the discovery and replication datasets were removed. Furthermore, dynamic *cis*-eQTL SNPs with pleiotropic effects significantly impacting more than one *cis*-eQTL gene that was causal of the outcome were also eliminated. The remaining genetic causal IVs were LD-clumped using function *ld_clump* in the R package *ieugwasr* (v0.1.5, Figure S9).

Finally, we conducted 2sGen-GPS on the causal dynamic *cis*-eQTLs in the combined dataset comprising three cohorts in Figure S8 and retained dynamic c*is*-eQTLs with a power ≥ 80% (Supplemental Methods). The same procedure was applied to 145,721 additive *cis*-eQTLs (representing 4,774 genes and 105,301 SNP IV predictors) with significance set at < 0.05/145,721 = 3.4×10^-7^. An importance test was conducted to prioritize the effect of all remaining causal *cis*-eQTLs on the longitudinal MDS-UPDRS scores using *PermutationImportance* in *eli5* package (v 0.13.0) in Python 3.8.3 (Supplemental Methods).

### Detecting shared temporal genetic effect between the longitudinal proteomics and PD progression

We applied 2sGen-GPS to detect shared temporal genetic effects between PD progression and longitudinal CSF proteomics, similar to the workflow as mentioned above. The proteomics data was generated using Olink Explore 1536 methodology. See the AMP-PD website (https://amp-pd.org/data/targeted-proteomics-data) for more details about the proteomics data. Initially, 2sGen-GPS was conducted on 1,472 CSF proteins, with the discovery dataset including 220 participants (136 HC and 84 PD cases) with 710 visits, and the replication dataset comprising 131 participants (58 HC and 99 PD cases) with 495 visits. The majority (95%) of participants in the CSF sample had fewer than five longitudinal time points. Normalized protein expression (NPX) was utilized to measure protein levels. Temporal causal *cis*-eQTLs associated with PD progression were utilized as predictors and adjusted for age at baseline, sex and PD diagnosis. Due to the limited sample size and time points available, the first-order lag was selected for the VAR model. We reported all temporal causal findings that passed a *P*-value threshold of 5.0×10^-5^ and with same direction of causal effect size in the discovery, replication and combine datasets.

### Detecting shared temporal genetic effect between the longitudinal neuroanatomic microstructural gradient features and PD progression

We conducted 2sGen-GPS to detect the shared temporal genetic effect between PD progression and change of voxel-level neuroanatomic microstructural gradient features following a workflow similar to the proteomics analysis, albeit without a replication dataset. Voxel-level neuroanatomic features were estimated with deformation-based morphometry (DBM)^33^ of T1-weighted MRI in the PPMI cohort, which included 171 PD cases with 559 visits. The follow-up time point of participants was fewer than four. The method for DBM and microstructural gradients features measurement^34^ are detailed in the Supplemental Methods. A total of 3,393 neuroanatomic microstructural gradient features were derived from 170 brain regions labeled by automated anatomical labelling atlas 3 (aal3)^35^ for 2sGen-GPS, and adjusted for image quality rating (IQR), age at disease onset, sex and disease duration years. A *P*-value threshold of 5.0×10^-5^ was considered significant.

### Temporal causal effect of genes on cognitive decline

We conducted 2sGen-GPS to detect the potential temporal causal effect of genes with temporal *cis*-eQTLs on the longitudinal MMSE^36^ score in patients with cognitive impairment (CI, Figure S12) following the procedure in Figure S9. The participants from MAP cohort within ROSMAP, featuring a longer follow-up period, was chosen as the discovery dataset, which comprises 409 CI cases (146 mild cognitive impairment (MCI) and 263 AD) with 3,543 visits. The replication dataset consisted of 563 CI cases (517 MCI and 46 AD) from the ADNI cohort, with 3,706 visits (Figure S12). Genotype data in the discovery dataset was imputed (Supplemental Methods, Figure S16), and the rsid and reference allele of SNPs in both the discovery and replication datasets were aligned with the findings of *cis*-eQTL SNPs. After this process, a total of 720 dynamic *cis*-eQTL SNPs for 392 genes (845 time-specific cis-eQTLs) and 114,463 additive cis-eQTL SNPs for 4,961 genes (150,806 additive cis-eQTLs) imputation were retained for the causality test after genetic IVs selection.

### Functional enrichment analysis and the connections between genes, drugs, and disease progression

GO enrichment analysis was performed on Metascape^37^. We compared the top 50 important causal genes of disease progression with gene expression changes caused by perturbations in the CMap^38^ database (from L1000). We focus on the perturbation compounds that may reverse gene expression in disease progression.

### Multidimensional tensor computation

In this study, we developed a tool *2sGen-GPS* based on multidimensional tensor computation (Figure S1) in the open-source general-purpose library, *PyTorch* (v1.7.0) for batch temporal and dynamic eQTL mapping and causality study. Additionally, other temporal eQTL and dynamic eQTL mapping approaches, including LR, Cubic, and AR(1), as well as MPTGA, are available within *2sGen-GPS*. The *2sGen-GPS* can be executed on CPUs or GPUs (see data and code availability).

## Results

### Temporal local genetic effects on the stationary longitudinal blood gene expression

The schematic of this study shown in Figure S1. We initially conducted temporal *cis*-eQTL mapping on 77,631,179 SNP-gene pairs (consisting of 17,222 stationary temporal genes expressed in blood and 5,951,191 SNPs, with SNP-gene distances < 1 Mb) in the discovery dataset (FDR < 0.05, 293 participants with 1,465 visits, Figure S2a-c), and then the significant signals were validated in an independent replication dataset (*P* < 0.05, including 346 participants with 1,730 visits, Figure S2d-f). We identified and replicated 774,533 significant SNP-gene pairs (representing 499,742 SNPs and 11,916 eGenes, Data 1) using MPTGA. It should be noted that these temporal *cis*-eQTLs were detected by testing the significance of the temporal genetic component on both genotype and the interaction between genotype and time (Methods). The replication rate of over 35% was observed in five temporal eQTL mapping approaches (Supplemental Methods, Figure S3a), and we found 11,268 eGenes with at least one temporal *cis*-eQTL from MPTGA can be detected by the other three temporal QTL approaches (Figure S3b, c).

### MPTGA in 2sGen-GPS identifies more dynamic genetic regulators of gene expression

We next carried out dynamic *cis*-eQTL mapping (Methods) by MPTGA, LR and Cubic approaches to dissect whether the temporal genetic regulatory patterns are varied over time. The total number of replicated dynamic *cis*-eQTLs was 6, 1,127, and 6,660 for LR, Cubic and MPTGA, respectively, with replication rates of 16.2%, 19.7%, and 29.5%, respectively (Figure 1a). We further compared the performance of the three dynamic eQTL mapping approaches using a simulated dataset with 18,000 longitudinal patterns, which were designed to mimic with and without dynamic eQTL effects (Supplemental Methods). The results indicated that the dynamic eQTL mapping approach in MPTGA outperformed LR and Cubic (*P* < 0.001,) across all auto-correlated data (Figure 1b), and the overall performance of the three approaches improved with higher auto-correlation (Figure 1b).

**Figure 1.**
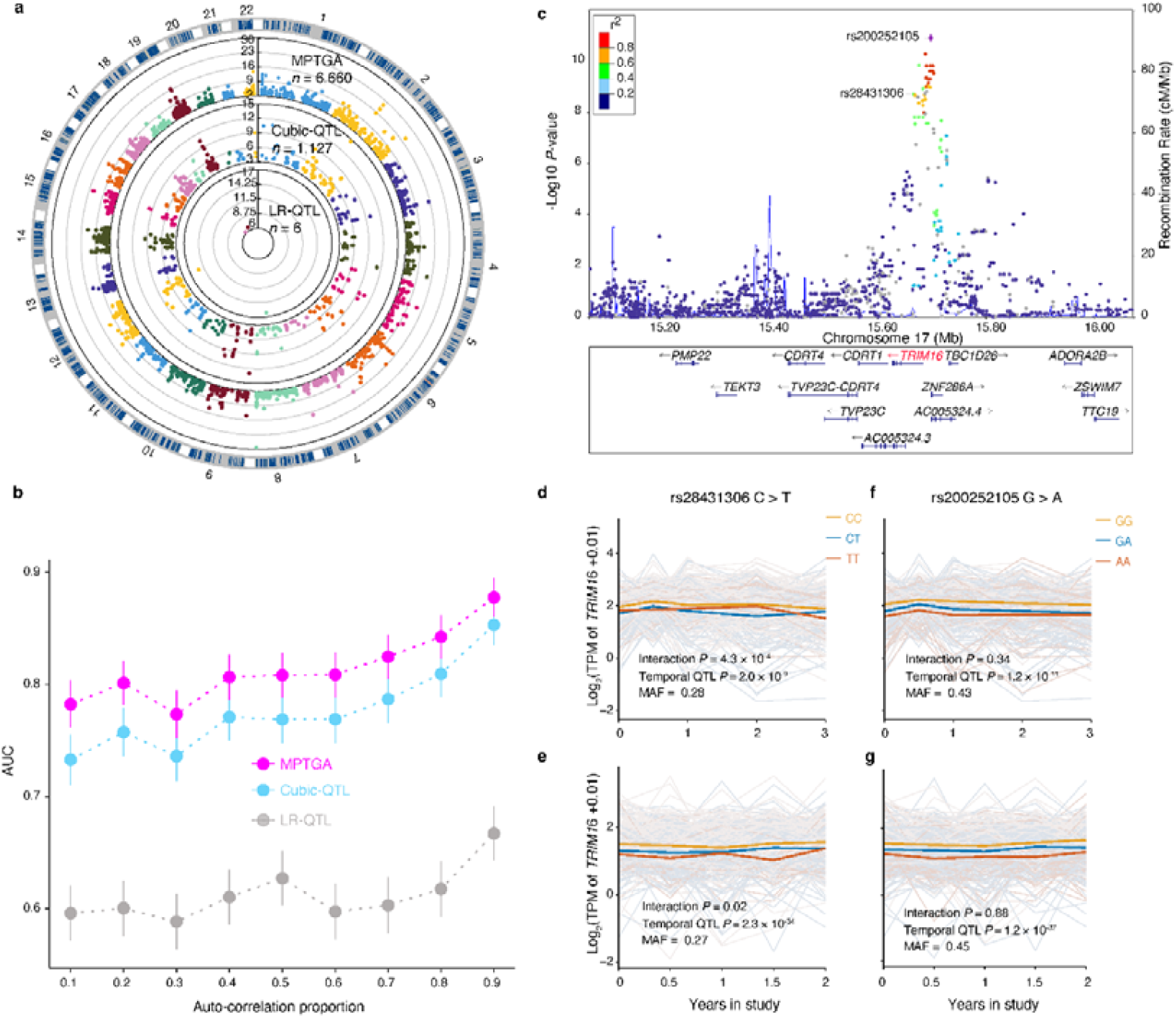
Identification of dynamic and additive *cis*-eQTLs. **a**, Circular Manhattan plot summarizing the results of significant dynamic *cis*-eQTLs in both discovery (interaction FDR < 0.05) and replication (raw interaction *P* value < 0.05) datasets using three regression based dynamic QTL mapping approaches. Inner track, linear regression (LR); middle track, cubic polynomial regression (Cubic); outer track, MPTGA. *n* is the number of dynamic *cis*-eQTLs. Values on the x axis are based on the eQTL SNP location, and values on the y axis are the -log10(interaction *P* value) in the discovery dataset. **b**, Area under the curve (AUC) of three regression based dynamic QTL mapping approaches under different strength of auto-correlation proportion in simulated dataset. All error bars indicate 95% CI. **c**, LocusZoom plot of the significant temporal *cis*-eQTL locus in gene *TRIM16*. Discovery (**d**) and replication (**e**) of dynamic *cis*-eQTL for *TRIM16*. Discovery (**f**) and replication (**g**) of additive *cis*-eQTL for *TRIM16*. Colors indicate SNP genotype, with orange as the minor allele; each translucent line represents the trajectory of *TRIM16* longitudinal expression level of a participant during the study; prominent lines represent the mean *TRIM16* longitudinal expression level through the study; interaction *P* value was determined for dynamic *cis*-eQTL, and temporal QTL *P* value was determined for temporal *cis*-eQTL using MPTGA.

By employing dynamic eQTL mapping in the MPTGA, 6,660 nonlinear dynamic *cis*-eQTLs (representing 5,438 SNPs and 1,563 eGenes) and 651,320 additive *cis*-eQTLs (representing 437,325 SNPs and 7,987 eGenes) were identified. The classifications of all the temporal eQTLs are provided in Data 1. Illustrative examples of temporal *cis*-eQTLs for the expression of *TRIM16* gene, indicating that a target gene with distinct genetic regulatory patterns (Figure 1c-g). Additional instances with varying effects of temporal *cis*-eQTLs on gene expression shown in Figure S5.

### Evaluating the temporal *cis*-eQTLs detected by MPTGA

We assessed the statistical power (Methods) for a random sample of 5,000 temporal *cis*-eQTLs from 774,533 significant SNP-gene pairs detected by MPTGA in the discovery dataset (with one side cutoff *α* = 0.001) and the replication dataset (with one side cutoff *α* = 0.05). There were over 4,000 temporal eQTLs exhibited statistical power exceeding 80% in both discovery and replication dataset (Figure S6b,c). Additionally, we evaluated the statistical power (Methods) for the same 5,000 temporal *cis*-eQTLs across sample sizes of 50, 100, 150, 200, 250, and 293 participants in the discovery dataset (with one side cutoff *α* = 0.001). We observed that only 3% of temporal eQTLs achieved 80% power with 50 participants and over 50% of temporal *cis*-eQTLs lacked statistical power when sample size less than 200 (Figure S6d).

Among the 774,533 significant temporal *cis*-eQTL, 49.1% of them were overlapped with the whole blood data from GTEx^15^ (Figure S7a). The majority of consistent eQTLs were additive, and only 7.6% of dynamic eQTLs were found in the GTEx (Figure S7a). Subsequently, we calculated the proportion of variance in phenotype explained (PVE) by each SNP for the temporal *cis*-eQTLs (Methods) in both the discovery and replication datasets. There was a strong concordance in PVE for each temporal *cis*-eQTL between the two datasets, with a Pearson correlation coefficient of 0.97 (Figure S7b). For 95% of the temporal *cis*-eQTLs, their PVE was less than 0.14 in both datasets (Figure S7b and Data 1), and more than 50% of the dynamic (Figure S7c, Table S3) and additive (Figure S7d, Table S4) *cis*-eQTL SNPs with PVE > 0.01 were located in the intronic region of gene.

### Identification of potential driver genes for the progression of PD

We employed 2sGen-GPS (Methods) to infer the temporally causal effect of genes on clinical phenotypes (MDS-UPDRS I-III^28,29^) linked to the progression of PD. The discovery dataset comprised 745 PD cases with 7,478 visits over a maximum of nine years from the PPMI cohort, while the replication dataset included 680 PD cases with 4,057 visits from the PDBP (follow up to five years) and SURE-PD3 (follow up to 2.5 years) cohorts (Figure S8). Both dynamic and additive *cis*-eQTL eGenes were selected for causality analysis. Initially, 650 dynamic *cis*-eQTL SNPs unaffected by candidate confounders (Supplemental Methods) were chosen as genetic instrument variables (IVs) for imputing the temporal genetic component of 406 longitudinal gene expressions (777 SNP-gene pairs). Employing 2sGen-GPS for longitudinal MDS-UPDRS II score (Figure S9), we identified and replicated one IV in a lncRNA gene (rs6578997-*AC051649*.*1*, Figure 2a,b, Data 2) that passed Bonferroni’s correction threshold (*P*□< □6.4×10^-5^, Figure S10a). For MDS-UPDRS III score, seven dynamic *cis*-eQTLs (Figure 2a, Data 2) including rs6578997-*AC051649*.*1* were detected. The result for a dynamic *cis*-eQTL, rs34660467-*NR2C1*, with third order lagged association with longitudinal MDS-UPDRS III is presented in Figure2c and Figure S10b. Subsequently, we conducted 2sGen-GPS on 145,721 additive *cis*-eQTLs (representing 4,774 genes and 105,301 IVs) for longitudinal phenotypes in PD. We uncovered and replicated 22 additive *cis*-eQTLs (representing 20 genes and 22 IVs) for MDS-UPDRS II and 499 additive *cis*-eQTLs (representing 411 genes and 499 IVs) for MDS-UPDRS III that pass Bonferroni’s correction threshold (*P*□< □3.4×10^-7^, Figure2a, Data 2). The result for an additive *cis*-eQTL, rs241437-*HLA-DRB5*, for longitudinal MDS-UPDRS II and III scores are presented in Figure2d and Figure S10c.

**Figure 2.**
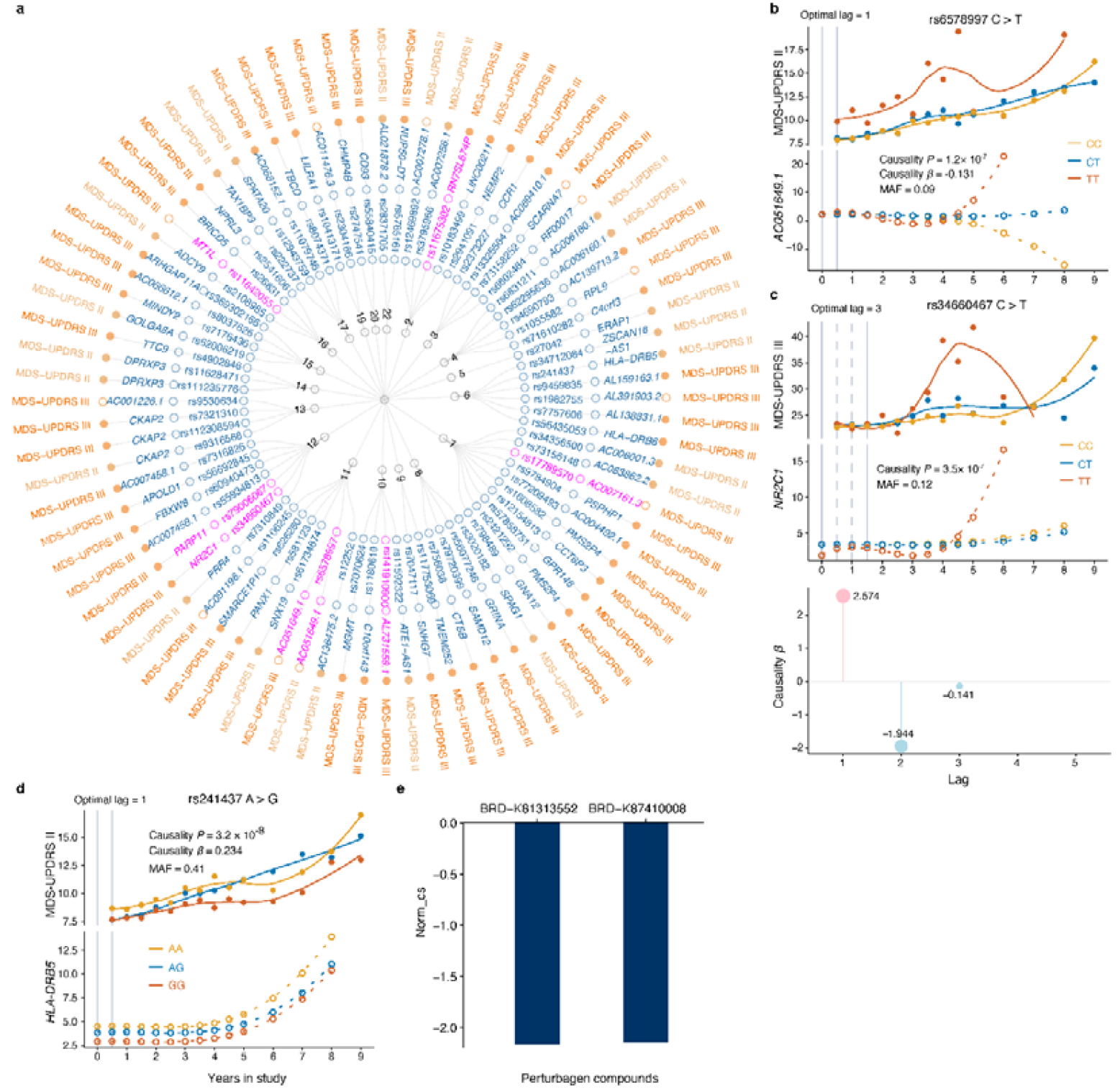
Temporal genetic causality analysis identifies dynamic driver genes for motor impairment progression in PD. **a**, Dendrogram shows the 80 independent causality test signals for two longitudinal clinical scores MDS-UPDRS II and III in PD. Moving outwards from the center are: (1) chromosome; (2) dynamic (magenta) or additive (blue) *cis*-eQTL SNPs; (3) dynamic (magenta) or additive (blue) *cis*-eQTL genes; (4) motor symptom clinimetric assessments in PD, yellow for MDS-UPDRS II and orange for MDS-UPDRS III. Outermost solid circles represent positive first order lag effect size of causal gene for phenotype while hollow circles represent negative effect size; notably, only top 50 prioritized additive *cis*-eQTL with causal effect on MDS-UPDRS III are presented. **b-d**, Visualization of the lagged correlation between nonlinear dynamic (*AC051649*.*1* and *NR2C1*) and additive (*HLA-DRB5*) *cis*-eQTL gene expression and motor impairment progression in the combined longitudinal PD dataset. Colors indicate SNP genotype, with orange being the minor allele. Dots and smooth lines in the top panel are the trajectory of mean longitudinal phenotype score and bottom panel shows the first order lagged temporal genetic component of causal gene expression during the study; the interval between two nearby grey vertical lines (solid or dashed) is the median first order lagged time in the time series of all participants; causality *P* value was determined for causality test for *cis*-eQTL gene to longitudinal phenotype level; causality *β* value was the lagged correlation effect sizes of gene on phenotype; notably, the causality *β* values of *NR2C1* on MDS-UPDRS III with 3rd order optimal lagged model were exhibited as a lollipop chart at the bottom of **c**, where dots indicate *β* of each order lagged time series of *NR2C1*; pink for positive effect and light bule for negative effect. **e**, Barplot shows the perturbagen compounds (x axis) and the connectivity score (y axis) computed by CMap which gene signatures connect (adjust *P* < 0.05) to the causal genes of PD progression. Norm_cs represents normalized connectivity score computed by CMap. Negative Norm_cs indicates the perturbagen compound may reverse the expression of driver genes.

In total, 525 temporal *cis*-eQTLs (427 genes and 525 IVs) causally associated with PD longitudinal motor phenotypes were identified. Gene Ontology (GO) enrichment analysis of the 427 blood causal genes of PD progression showed that multiple biological processes were immune response (Figure S10d). Among the 427 causal genes, 44 have been implicated in PD according to the *DisGeNET*^39^ platform (UMLS CUI: C0030567). We also prioritized the contribution of these causal temporal *cis*-eQTLs on the longitudinal MDS-UPDRS II and III scores, respectively (Figure 2a and Data 2). In the optimal lag for 525 IV-gene-phenotype VAR models (Methods) for the longitudinal phenotypes during the progression of PD, we observed 119 models with a single time lag, 128 models with two time lags, 194 models with three time lags and only 11 models with five time lags (Data 2), where the median interval time of a lag in the PD datasets was 0.5 year (Figure 2b-d, Figure S10a-c). The optimal lag, such as 1 or 3 in Figure 2b, c, indicates that the lagged correlation effect of the causal genes on the phenotype may have appeared at least half or one year and a half ago. The VAR model with higher-order lag suggests a more complex perturbed relationship between the genetic component of gene expression and phenotype (Figure 2c).

We further connected the top 50 important causal genes of PD progression with gene expression changes caused by perturbations in the CMap database (from L1000). There were two small molecule compounds (BRD-K81313552 and BRD-K8741008) may reverse gene expression in PD progression and can be potentially repurposed to slowing or halting the progression of disease (Figure 2e).

### Cerebrospinal fluid proteomic mediators link peripheral genomic drivers to Parkinson’s disease progression

While our longitudinal analysis identified dynamic associations between peripheral gene expression and Parkinson’s disease (PD) motor progression, the extent to which these peripheral factors infiltrate the central nervous system (CNS) to drive disease pathology remains to be elucidated. Given the near-total lack of longitudinal human brain tissue, we utilized cerebrospinal fluid (CSF) proteomics as a proxy to investigate shared temporal genetic effects between circulating signals and PD progression (Methods). By integrating 525 temporal *cis*-eQTLs causal for PD progression with the longitudinal expression profiles of 1,472 CSF proteins, we identified 47 IV-gene-protein trios (comprising 30 IVs, 29 eGenes, and 44 proteins; Table S3) via Vector Autoregression (VAR) modeling (*P* < 5×10^-5^), which were subsequently validated (*P* < 0.05). Notably, nine of these CSF proteins have established clinical associations with PD according to the *DisGeNET* database (Figure 3a).

**Figure 3.**
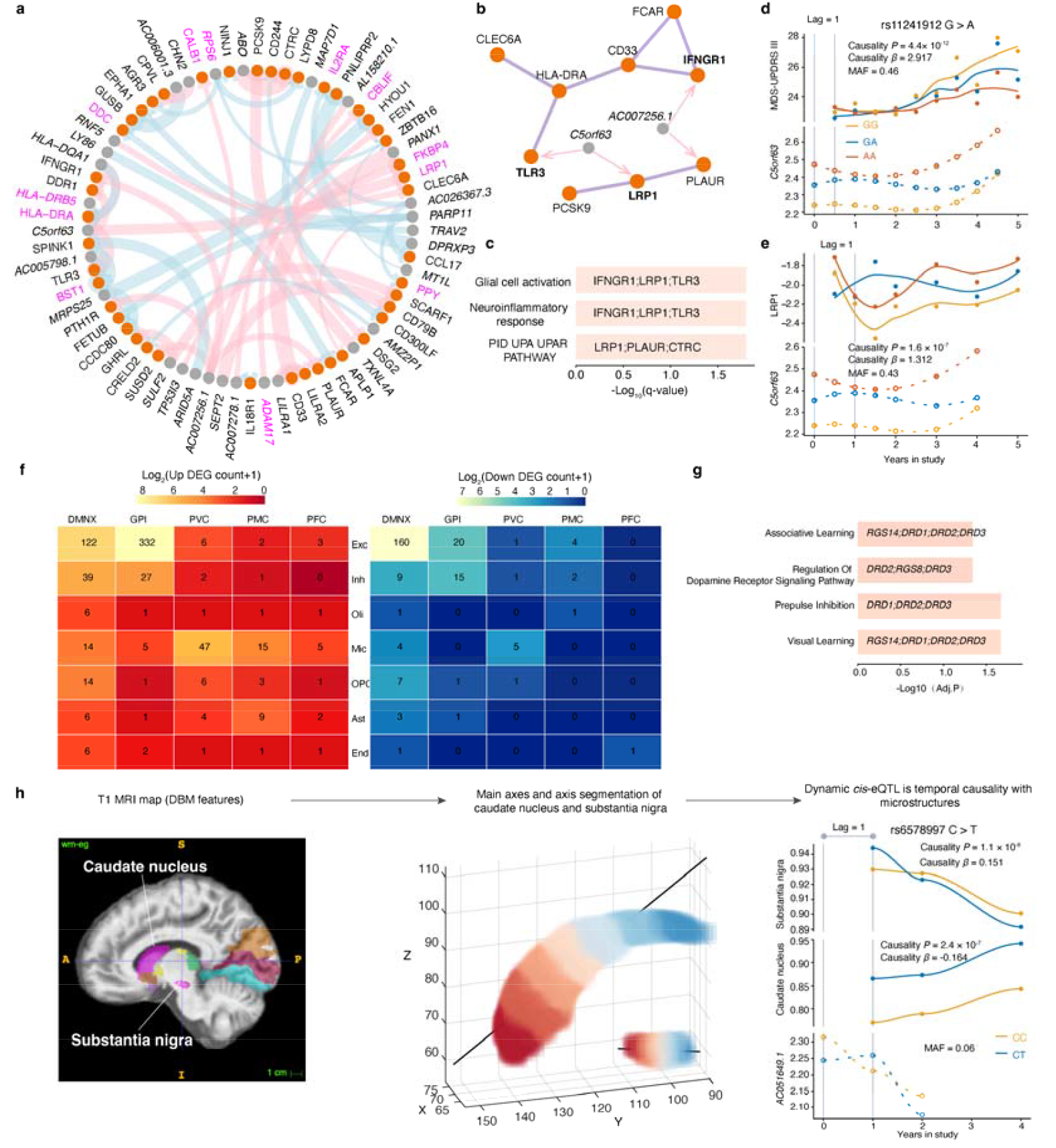
Shared temporal genetic effects among CSF proteins, neuroanatomic features and PD motor symptoms varying over time. **a**, Circle plot shows the 29 temporal *cis*-eQTL genes (grey node) which are causal of PD progression with causal effects on 44 longitudinal targeted CSF proteins (orange node). The edge thickness quantifies the strength of causal effect of gene on the protein, and the edge colors indicate direction of effect size, with positive (pink) or negative (light blue) effect. The texts with magenta indicate the genes or proteins have been reported to associated with PD. Protein-protein interactions (**b**) among 44 CSF proteins reveal the involvement of three CSF proteins (bold text) in connecting to glial cell activation or neuroinflammatory response (**c**). Two grey nodes indicate two causality blood genes of PD progression that are positive lagged association with CSF proteins. Gene *C5orf63* is temporal causality with PD motor symptom (**d**) and CSF protein LRP1 (**e**). Participants in **d** using the same dataset (PPMI, PDBP and SURE-PD3) in Figure 2b-d; participants in **e** using the combined longitudinal dataset (PPMI and PDBP, Methods). **f**, Heatmaps illustrating the number of significantly up-regulated (left) and down-regulated (right) differentially expressed genes (DEGs) in carriers of the rs11241912 mutation compared to non-carriers. Analysis was conducted across five distinct brain regions: Dorsal Motor Nucleus of the Vagus (DMNX), Globus Pallidus Internus (GPI), Premotor Cortex (PMC), Prefrontal Cortex (PFC), and Primary Visual Cortex (PVC). Cell types include astrocytes (Ast), excitatory neurons (Exc), microglia (Mic), inhibitory neurons (Inh), oligodendrocytes (Oli), oligodendrocyte progenitor cells (OPC) and endotheliocyte (End). Significance was defined at a False Discovery Rate (FDR) < 0.05. Numbers within each cell indicate the total count of DEGs exhibiting an absolute log fold change > 1 for each region–cell type pair. **g**, Horizontal bar plot illustrating the top four enriched GO biological processes for genes up-regulated in GPI Exc of rs11241912 carriers. Statistical significance is expressed as -log10 (FDR) (Fisher’s exact test). Specific core enrichment genes are annotated within each bar. **h**, LncRNA *AC051649*.*1* that is temporal causality with PD motor symptom may be causal of substantia nigra and caudate nucleus varying over time in PD. Colors in the brain MRI image (left) along the Anterior to Posterior (AP) axis indicate 29 candidate brain region that may be affected by the *AC051649*.*1* expression varying over time; visualization of the DBM microstructural gradients along the AP axis of the caudate nucleus and substantia nigra on a T1-weighted image of a sample (middle); temporal association between *AC051649*.*1* and left caudate nucleus 7 and left substantia nigra pars reticulata 3 in PC1 (right).

Protein-protein interaction (PPI) and Gene Ontology (GO) enrichment analyses of the 44 candidate CSF proteins highlighted a functional cluster involved in glial cell activation and neuroinflammatory responses (Figure 3b,c). Specifically, we found that the temporal genetic component of *C5orf63*, regulated by the *cis*-variant rs11241912, exhibited a significant positive lagged correlation with both MDS-UPDRS III scores (*β* = 2.917) and CSF levels of LRP1 (*β* = 1.312) across discovery and replication datasets (Figure 3d,e; Figure S11a,b). Given that LRP1 is a recognized receptor for α-synuclein uptake and propagation, these results suggest a specific peripheral-to-central regulatory axis.^40^.

To resolve the neuroanatomical and cellular susceptibility associated with rs11241912, we interrogated a cross-sectional, multi-region single-cell transcriptomic atlas (Supplemental Methods). We identified highly localized regulatory signatures within the dorsal motor nucleus of the vagus (DMNX) and the globus pallidus internus (GPI) - two regions critically involved in early-stage PD pathology (Figure 3f). Notably, excitatory (Exc) neurons in the GPI of rs11241912 carriers displayed a profound transcriptional shift, characterized by a predominant up-regulation of genes (332 up-regulated vs. 20 down-regulated DEGs). Functional enrichment of these up-regulated DEGs revealed a robust over-representation of the “Regulation of Dopamine Receptor Signaling Pathway”, driven by core genes including *DRD2, RGS8*, and *DRD3* (Figure 3g). Collectively, these data suggest that the rs11241912-mediated peripheral signal may influence motor progression by modulating dopamine receptor sensitivity and proteostatic receptors like LRP1 within the GPI circuitry.

### Shared temporal genetic effects between neuroanatomic features and PD progression

Furthermore, we applied 2sGen-GPS to analyze 3,393 temporal voxel-level neuroanatomic microstructural gradient features in a PD dataset (Methods). This analysis identified 456 IV-gene-neuroanatomic VAR models (representing 24 IVs, 24 genes and 266 neuroanatomic features) that passed Bonferroni’s correction (*P*□< □5×10^-5^, Table S4). Notably, the lncRNA *AC051649*.*1* gene, dynamically regulated by the rs6578997 variant *in cis*, was found to be causal of 159 neuroanatomic microstructural gradient features in 29 brain regions, including 58 substantia nigra features and seven caudate nucleus features (Figure 3h, Table S4).

### Identification of temporal *cis*-eQTLs as potential driver genes for AD cognitive decline

We extended 2sGen-GPS (Methods) to infer the temporally causal effect of genes on longitudinal mini-mental state examination (MMSE)^36^ score linked to the cognitive impairment (CI) in two independent AD cohorts, both of them with more than 15 years follow-up visits. Specifically, we employed a total of 409 CI cases with 3,543 visits from the MAP cohort as the discovery dataset, and the remaining 563 CI cases with 3,706 visits from the ADNI cohort (Figure S12) were designated as the replication dataset. A total of 720 dynamic and 114,463 additive *cis*-eQTL SNPs, unassociated with candidate confounders (Supplemental Methods) were selected as genetic IVs for the imputation of the temporal genetic component of 5,353 longitudinal gene expressions (845 dynamic and 150,806 additive *cis*-eQTLs). Employing 2sGen-GPS for the longitudinal MMSE score, following LD clumping and power estimation (Figure S9, Supplemental Methods), we identified and replicated 13 dynamic (representing 13 genes and 13 IVs, *P*□< □0.05/845 = 5.9×10^-5^) and 396 additive (representing 368 genes and 396 IVs, *P*□< □0.05/150,806=3.3×10^-7^) *cis*-eQTLs that passed Bonferroni’s correction (Data 2). In the context of the optimal lag in these IV-gene-trait VAR models, we observed 402 models with a single time lag, six models with two time lags, and only one model with three time lags (Data 2). The median interval time for a lag in the CI datasets was one year.

Among the 381 causal genes, 53 genes have been linked to AD or CI, as reported in the *DisGeNET* platform (UMLS CUI: C0002395 and C0338656). PIP GO enrichment analysis of the 381 blood causal genes of cognitive decline showed that the most significant biological process was response to oxidative stress (Figure S13a). For instance, the *SNCA* gene was dynamically regulated by the rs17016168 variant in *cis* and exhibited a lagged negative correlation with the longitudinal MMSE score (Figure 4a, b, Figure S13b). *SNCA* has been proposed as a shared genetic etiology with AD and PD^41^. We next prioritized the contribution of these 381 causal genes with respect to the longitudinal MMSE score. Notably, the *RN7SL674P* gene, dynamically regulated by rs11675302 variant in *cis*, was the highest prioritized compared to other dynamic eQTL genes (Figure 4a, c, Figure S13c, Data 2).

**Figure 4.**
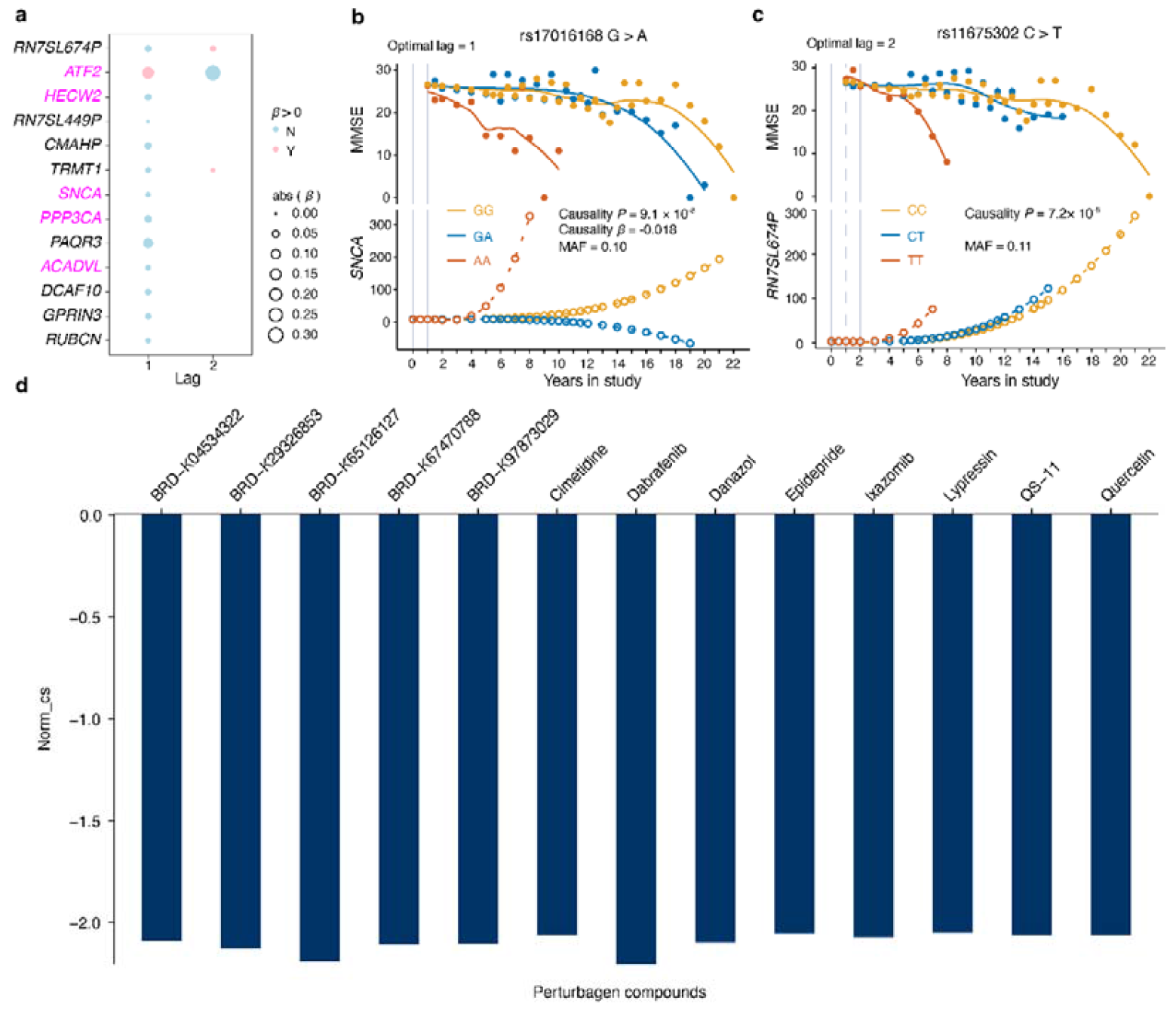
Temporal genetic causality analysis identifies dynamic driver genes for cognitive decline. **a**, Dot plot shows the 13 prioritized dynamic *cis*-eQTL genes which are causal of cognitive decline measured by longitudinal MMSE score in AD cohorts. The x axis is the lag in an optimal temporal causality model for a dynamic *cis*-eQTL gene, and the dot size quantifies the strength of lagged correlation of gene on the MMSE score varying over time. Dot colors indicate direction of lagged correlation at each lag, with positive (plink) or negative effect (light blue). The texts with magenta indicate the genes have been reported to associated with AD or CI. **b, c**, Visualization of lagged correlation between dynamic *cis*-eQTL gene expressions (*SNCA* and *RN7SL674P*) and longitudinal MMSE linked to cognitive decline in the combined AD longitudinal dataset. **d**, Barplot shows the perturbagen compounds (x axis) and the connectivity score (y axis) computed by CMap which gene signatures connect (adjust *P* < 0.05) to the causal genes of cognitive decline. Norm_cs represents normalized connectivity score computed by CMap. Negative Norm_cs indicates the perturbagen may reverse expression of disease genes.

We connected the top 50 important causal genes of cognitive decline with gene expression changes caused by perturbations in the CMap database (from L1000). We found 13 small molecule compounds (Figure 4d, Table S5) may reverse expression of causal genes in cognitive decline. Notably, quercetin has been extensively studied for AD treatment^42^.

### Discussions

In this study, we developed 2sGen-GPS, a robust analytical framework designed to systematically decode the functional consequences of genetic variants and genes driving neurodegenerative progression. By applying the MPTGA component of our framework to two independent PD cohorts, we identified an extensive repertoire of temporal *cis*-eQTLs, including 6,660 nonlinear dynamic and 651,320 additive regulatory signals. Our results underscore the inherent limitations of conventional “snapshot” analyses, which frequently fail to capture the transient and evolving nature of dynamic *cis*-eQTLs (Figure S7a). Through both simulated and real-world data validation, 2sGen-GPS demonstrated superior accuracy and sensitivity in detecting these temporal signatures compared to existing methodologies (Figure 1a,b).

Beyond methodological innovation, the application of 2sGen-GPS to neurodegenerative disease cohorts uncovered a multitude of novel dynamic driver genes linked to motor impairment in PD and cognitive decline in AD. Central to these findings is the identification of a peripheral-to-central regulatory axis—most notably involving the rs11241912-driven expression of *C5orf63* - which modulates PD progression through CSF-mediated signals and cell-type-specific responses in the GPI. Crucially, our temporal causality analysis reveals a significant proactive regulatory window: the majority of identified driver genes begin to influence motor impairment at least six to eighteen months prior to clinical manifestation, with a similar one-year lead time observed for cognitive impairment in AD. These findings highlight the potential for 2sGen-GPS to identify early-warning molecular markers and nominate time-sensitive therapeutic targets long before irreversible clinical decline occurs.

Our study identifies a novel peripheral-to-central regulatory axis linked to the rs11241912 variant. The positive lagged correlation between peripheral *C5orf63* expression and CSF LRP1 levels suggests a systemic reinforcement of central proteostasis. LRP1 has been identified as a critical neuronal receptor for the uptake and spread of α-synuclein^40^. We propose that the rs11241912-driven elevation of LRP1 facilitates the efficient sequestration or clearance of extracellular α-synuclein, thereby mitigating neurotoxicity. This molecular resilience is concentrated in GPI excitatory neurons, where rs11241912 carriers exhibit a profound up-regulation of the dopamine receptor signaling pathway (*DRD2, RGS8* and *DRD3*). Rather than a simple change in cell number, this transcriptional reprogramming likely potentiates the responsiveness of the GPI circuitry to residual dopamine, explaining the lower MDS-UPDRS III scores observed in carriers. However, this compensatory endotype is transient; its eventual exhaustion (Figure S11c) leads to the resumption of clinical decline, defining a critical therapeutic window for disease intervention.

We also uncovered a series of genes that exhibit shared temporal genetic effects between neuroanatomic features and motor impairment progression in PD (Figure 3h, Table S4), which linked the potential role of peripheral driver on the CNS. We identified a dynamic *cis*-eQTL gene, *AC051649*.*1*, being negative lagged correlation with both longitudinal MDS-UPDRS II and III. Interestingly, we observed that the *AC051649*.*1* was positive and negative lagged correlation with the substantia nigra and caudate nucleus microstructural gradient features in the progression of PD (Figure 3h, Table S4), respectively. The atrophy of the substantia nigra during PD progression is consistent with previous study^43,44^. Typically, the volume of the caudate nucleus in PD case is lower than in healthy controls^34,45^, while we observed an enlargement of the caudate nucleus during the study. The caudate nucleus is part of the striatum and compensatory mechanisms of increase in striatal dopamine turnover has been shown to occur in early PD ^46,47^. Whether the *AC051649*.*1* gene, regulated by rs6578997 variant, is causal of enlargement of the caudate nucleus and associated with compensatory mechanisms of striatal dopamine in early PD progression warrants further study.

Compelling evidence across diverse studies highlights oxidative stress as a fundamental driver of Alzheimer’s disease (AD) pathogenesis and its subsequent clinical progression^48^. Through our 2sGen-GPS framework, we identified twelve genes (*BAK1, CAT, ATF2, ERRCC3, GPX7, KPT1, SNCA, PRDX6, PRDX5, ERO1A, RBM11* and *SRXN1*) that are functionally linked to the cellular response to oxidative stress (GO:0006979) and exert a temporal causal influence on cognitive decline. These genetic signatures provide a molecular rationale for antioxidant-based interventions.^42,49^. Our drug repurposing analysis suggested quercetin (Figure 4d), a flavonols class drug, as a potential drug for slowing or halting the progression of cognitive impairment. Cimetidine, which may enhance the antioxidant defense mechanism^50^, is another suggested drug (Figure 4d) for cognitive decline. However, a pharmaco-epidemiological cohort study reported that cimetidine histamine 2 receptor antagonists (H2RAs) treatment increased the risk of dementia^51^. Consequently, the pharmacological efficacy and safety profiles of the 13 prioritized compounds (Figure 4d) warrant rigorous longitudinal validation in clinical settings to determine their true potential in halting neurodegenerative progression.

In this study, the temporal and dynamic eQTL analysis involved a larger sample size and more follow-up visits (3,195 follow-up samples across five time points) compared to a previous study (232 follow-up samples across two time points) investigating temporal genetic effects on blood gene expression in aging^52^. Our 2sGen-GPS methods offer several improvements over existing approaches. The MPTGA in 2sGen-GPS is extended from two temporal eQTL mapping approaches, Funmap2^21,53^ and MPTGA in Lin et. al.^22^, which are failing to addressing the effect of confounding factors. For longitudinal cohort studies, numerous confounders, such as age, sex and population structure as well as latent unknown factors for gene expression, may contribute to variations in temporal QTL mapping. For MPTGA, although the regularization term may limit sensitivity and increase computational complexity for identifying temporal eQTLs ( Supplemental Methods, Supplemental Discussion, Figure S14), the assumption^21,22,54^ of longitudinal gene expression following a multivariate normal density and related variance at each time point makes MPTGA more sensitive and accurate in capturing dynamic effects to identify dynamic eQTLs (Figure 1a, b). Granger causality test has been widely used in inferring the association between two temporal traits in biomedical studies^22,27,55,56^, but the risk of spurious correlation occur in the lagged correlation in Granger ‘causality’^26,27^. In the context of 2sGen-GPS, similar to two stage MR^8,25^ analysis framework, we rigorously used genetic IV from the temporal *cis*-eQTL SNPs for longitudinal gene expression imputation (Methods) to enhance causal inferences on lagged correlation analysis. Prediction on the genetic component of expression could avoid confounding influences on causality outcomes^13^, making it applicable for diverse time courses in population (Figure 2,4). Lin et.al.^22^ indicated that at least eight time points and 200 individuals were needed for the effective application of causality tests in a human study. Considering the confounders, the number of time point and sample size in our dataset is sufficient to detect temporal causality associations using 2sGen-GPS. Furthermore, comparing with a previous temporal genetic causality test approach developed by Lin et.al.^22^, using VAR model for Granger causality testing^26,27^ allowed us to infer Granger causal connections at longer lags at individual level (Data 2) on the dataset comprising multiple time series in numerous participants.

Several limitations of this study must be considered. The MPTGA in 2sGen-GPS utilized a cubic polynomial model as the basic function to capture nonlinear dynamic patterns, and did not apply higher degree polynomials to fit the gene expression dataset with the limited longitudinal visits in this study. Previous research has demonstrated that a cubic polynomial model is sufficient to capture dynamic patterns in time series data with six time points^22^. Another limitation of MPTGA is the assumption that variances at each time point follow an AR(1) model. A more generalized assumption of variances, tailored to specific characteristics of data could potentially enhance the accuracy of our QTL approach in the context of longer and more complex longitudinal datasets^53^. In the context of eQTL linear regression analysis using ‘snapshot’ molecular data with a minor allele frequency (MAF) threshold of 0.05, a minimum sample size of approximately 70 samples is preferred^57^. While the power of the majority of temporal eQTLs in our MPTGA findings was sufficient, as evidenced by the inclusion of 293 and 346 participants with five visits in both the discovery and replication datasets, there were still a subset of temporal eQTLs that were underpowered (Figure S6b, c). These weaknesses could be better overcome as larger sample size with longer visit of gene expression data become available. In addition, our 2sGen-GPS only includes one IV-gene predictor, and combining *cis*-SNPs into a single predictor and integrating diverse predictors may capture heterogeneous signals better than individual *cis*-eQTLs^7,13^. Our approach can be updated using causal genes-disease network to adapt to the complexity of genetic regulation. Another caveat pertain to our findings is casual interpretation. The models of longitudinal gene expression imputation for causality tests were trained from a dataset with three years following-up visits, while the study years of dependent variables extend beyond three years (Figure S8a, Figure S12a). Although we observed that the PVE of these IVs is concordance between discovery and replication datasets, there is a lack of additional data to test the accuracy of gene expression prediction over longer periods. Furthermore, the predicted gene models were trained in a dataset comprising healthy controls and PD cases, and longitudinal gene expression data in AD is needed to ascertain whether these temporal eQTLs’ regulatory patterns are similar in the context of AD.

### Conclusions

We present 2sGen-GPS approach to better understand the causal genetic variants that account for the heterogeneity of progression in neurodegenerative diseases. Our MPTGA in 2sGen-GPS exhibits more accurate and sensitive in identifying dynamic *cis*-eQTLs. The temporal eQTLs and probable causal genes will pave the way for offering new insights into the underlying mechanisms of genetic basis of progression of PD and cognitive decline in AD. These findings not only enhance our understanding of the dynamic genetic effects on progression of disease, but also aid in achieving the goals of precision medicine for neurodegenerative diseases. With the advent of longitudinal omics or imaging data in cohort study, we believe that it is time for temporal QTL to receive a similar degree of support to their snapshot QTL counterparts, and we anticipate that this analysis framework will be indispensable for aging-associated cohort studies integrating genetics and phenotypes in the foreseeable future.

## Data Availability

The longitudinal RNA sequencing TPM data for PPMI and PDBP included in this study are publicly available upon request to AMP PD (https://amp-pd.org/) through a Data Agreement. WGS data and clinical data for PPMI, PDBP and SURE-PD3 is also available through AMP PD (https://amp-pd.org/). Brain image data for PPMI is available through. Longitudinal clinical data for ROSMAP resources can be requested at https://www.radc.rush.edu. ROSMAP genotype array raw data is available via the AD Knowledge Portal (https://adknowledgeportal.org). The AD Knowledge Portal is a platform for accessing data, analyses, and tools generated by the Accelerating Medicines Partnership (AMP-AD) Target Discovery Program and other National Institute on Aging (NIA)-supported programs to enable open-science practices and accelerate translational learning. The data, analyses and tools are shared early in the research cycle without a publication embargo on secondary use. Data is available for general research use according to the following requirements for data access and data attribution (https://adknowledgeportal.synapse.org/Data%20Access). For access to content described in this manuscript see: https://doi.org/10.7303/syn2580853. Longitudinal clinical data and WGS data for ADNI are available through https://adni.loni.usc.edu.

https://www.radc.rush.edu

https://adni.loni.usc.edu

## Data and code availability

The analysis pipeline code is available at *https://github.com/sixguns1984/2sGen-GPS*. The longitudinal RNA sequencing TPM data for PPMI and PDBP included in this study are publicly available upon request to AMP PD (*https://amp-pd.org/*) through a Data Agreement. WGS data and clinical data for PPMI, PDBP and SURE-PD3 is also available through AMP PD (*https://amp-pd.org/*). Brain image data for PPMI is available through. Longitudinal clinical data for ROSMAP resources can be requested at *https://www.radc.rush.edu*. ROSMAP genotype array raw data is available via the AD Knowledge Portal (https://adknowledgeportal.org). The AD Knowledge Portal is a platform for accessing data, analyses, and tools generated by the Accelerating Medicines Partnership (AMP-AD) Target Discovery Program and other National Institute on Aging (NIA)-supported programs to enable open-science practices and accelerate translational learning. The data, analyses and tools are shared early in the research cycle without a publication embargo on secondary use. Data is available for general research use according to the following requirements for data access and data attribution (https://adknowledgeportal.synapse.org/Data%20Access). For access to content described in this manuscript see: https://doi.org/10.7303/syn2580853. Longitudinal clinical data and WGS data for ADNI are available through *https://adni.loni.usc.edu*.

## Acknowledgements

G.L.’s work is supported by National Natural Science Foundation of China (32470708, 32270701), Shenzhen Fundamental Research Program (JCYJ20240813151132042), Shenzhen Key Laboratory for Systems Medicine in Inflammatory Diseases (ZDSYS20220606100803007) and The Science and Technology Planning Project of Guangdong Province (2023B1212060018). J.L.’s work is supported by Sun Yat-sen Pilot Scientific Research Fund (YXQH202516). This work also supported by High-performance Computing Public Platform (Shenzhen Campus) of SUN YAT-SEN UNIVERSITY.

We thank all study participants and their families, investigators and members of the following knowledge platform: Accelerating Medicine Partnership Parkinson’s Disease (AMP PD), and studies: Parkinson’s Progression Markers Initiative (PPMI); Parkinson’s Disease Biomarker Program (PDBP); Study of Urate Elevation in Parkinson’s Disease, phase 3 (SURE-PD3); Accelerating Medicines Partnership in Alzheimer’s Disease (AMP-AD); Religious Orders Study and Memory and Aging Project (ROSMAP); Alzheimer’s Disease Neuroimaging Initiative (ADNI). **AMP PD**: Data used in the preparation of this article were obtained from the Accelerating Medicine Partnership® (AMP®) Parkinson’s Disease (AMP PD) Knowledge Platform. For up-to-date information on the study, visit https://www.amp-pd.org.

The AMP® PD program is a public-private partnership managed by the Foundation for the National Institutes of Health and funded by the National Institute of Neurological Disorders and Stroke (NINDS) in partnership with the Aligning Science Across Parkinson’s (ASAP) initiative; Celgene Corporation, a subsidiary of Bristol-Myers Squibb Company; GlaxoSmithKline plc (GSK); The Michael J. Fox Foundation for Parkinson’s Research ; Pfizer Inc.; AbbVie Inc.; Sanofi US Services Inc.; and Verily Life Sciences.

ACCELERATING MEDICINES PARTNERSHIP and AMP are registered service marks of the U.S. Department of Health and Human Services.

**PPMI**: Data used in the preparation of this article were obtained [on November, 14 2022] from the Parkinson’s Progression Markers Initiative (PPMI) database (www.ppmi-info.org/access-dataspecimens/download-data), RRID:SCR 006431. This analysis used MRI imaging data for PD participants, obtained from PPMI upon request after approval by the PPMI Data Access Committee. For up-to-date information on the study, visit www.ppmi-info.org. PPMI – a public-private partnership – is funded by the Michael J. Fox Foundation for Parkinson’s Research and funding partners, including 4D Pharma, Abbvie, AcureX, Allergan, Amathus Therapeutics, Aligning Science Across Parkinson’s, AskBio, Avid Radiopharmaceuticals, BIAL, Biogen, Biohaven, BioLegend, BlueRock Therapeutics, Bristol-Myers Squibb, Calico Labs, Celgene, Cerevel Therapeutics, Coave Therapeutics, DaCapo Brainscience, Denali, Edmond J. Safra Foundation, Eli Lilly, Gain Therapeutics, GE HealthCare, Genentech, GSK, Golub Capital, Handl Therapeutics, Insitro, Janssen Neuroscience, Lundbeck, Merck, Meso Scale Discovery, Mission Therapeutics, Neurocrine Biosciences, Pfizer, Piramal, Prevail Therapeutics, Roche, Sanofi, Servier, Sun Pharma Advanced Research Company, Takeda, Teva, UCB, Vanqua Bio, Verily, Voyager Therapeutics, the Weston Family Foundation and Yumanity Therapeutics.

**PDBP**: Parkinson’s Disease Biomarker Program (PDBP) consortium is supported by the National Institute of Neurological Disorders and Stroke (NINDS) at the National Institutes of Health. A full list of PDBP investigators can be found at https://pdbp.ninds.nih.gov/policy. The PDBP Investigators have not participated in reviewing the data analysis or content of the manuscript.

**SURE-PD3:** The Study of Urate Elevation in Parkinson’s Disease, Phase 3 (SURE-PD3) is funded by the National Institute of Neurological Disorders and Stroke (NINDS) at the National Institutes of Health with support from The Michael J. Fox Foundation and the Parkinson Study Group. For additional study information, visit https://clinicaltrials.gov/ct2/show/NCT02642393. The SURE-PD3 investigators have not participated in reviewing the data analysis or content of the manuscript.

**AMP AD:** The results published here are in whole or in part based on data obtained from the AD Knowledge Portal.

**ROSMAP:** We thank the study participants and staff of the Rush Alzheimer’s Disease Center. ROSMAP is supported by P30AG10161, P30AG72975, R01AG15819, R01AG17917. U01AG46152, and U01AG61356.

**ADNI:** Data collection and sharing for this project was funded by the Alzheimer’s Disease Neuroimaging Initiative (ADNI) (National Institutes of Health Grant U01 AG024904) and DOD ADNI (Department of Defense award number W81XWH-12-2-0012). ADNI is funded by the National Institute on Aging, the National Institute of Biomedical Imaging and Bioengineering, and through generous contributions from the following: AbbVie, Alzheimer’s Association; Alzheimer’s Drug Discovery Foundation; Araclon Biotech; BioClinica, Inc.; Biogen; Bristol-Myers Squibb Company; CereSpir, Inc.; Cogstate; Eisai Inc.; Elan Pharmaceuticals, Inc.; Eli Lilly and Company; EuroImmun; F. Hoffmann-La Roche Ltd and its affiliated company Genentech, Inc.; Fujirebio; GE Healthcare; IXICO Ltd.; Janssen Alzheimer Immunotherapy Research & Development, LLC.; Johnson & Johnson Pharmaceutical Research & Development LLC.; Lumosity; Lundbeck; Merck & Co., Inc.; Meso Scale Diagnostics, LLC.; NeuroRx Research; Neurotrack Technologies; Novartis Pharmaceuticals Corporation; Pfizer Inc.; Piramal Imaging; Servier; Takeda Pharmaceutical Company; and Transition Therapeutics. The Canadian Institutes of Health Research is providing funds to support ADNI clinical sites in Canada. Private sector contributions are facilitated by the Foundation for the National Institutes of Health (www.fnih.org). The grantee organization is the Northern California Institute for Research and Education, and the study is coordinated by the Alzheimer’s Therapeutic Research Institute at the University of Southern California. ADNI data are disseminated by the Laboratory for Neuro Imaging at the University of Southern California.

## Author contributions

G.L. conceived and designed the study. J.L. contributed to the study design, and carried out the statistical and bioinformatics analyses. Data were collected by J.L., H.W. and W.D. G.L. and J.L. drafted the manuscript. All authors reviewed, edited and approved the manuscript prior to submission.

