## Supplemental Information for "Nonlinear dynamic genetic regulation identifies peripheral drivers of neurodegenerative disease progression"

Nonlinear dynamic genetic regulation reveals putative drivers for the progression in neurodegenerative diseases

[Data 1. Blood temporal *cis*-eQTLs at FDR<0.05 comprising nonlinear dynamic and additive *cis*-eQTLs .	36](#_Toc220588131)

### Supplemental Discussion

#### Evaluation of temporal eQTL mapping approaches

Applied to two longitudinal RNA-seq datasets from human blood, the MPTGA in 2sGen-GPS exhibited better replication rate than other three time series models in identifying temporal eQTLs (Figure S3a), but not Union. Union, as a greedy method in temporal eQTL mapping, only requires a significance at any time point (Methods), potentially resulting in a higher false positive rate compared to MPTGA (Figure S14b). Furthermore, Union is unable to capture dynamic information^1,2^. The basis function of MPTGA in 2sGen-GPS extends from MPTGA ^2^ and is similar to the Cubic method, with the addition of correction terms and a rederived equation (Methods and Supplemental Methods) to adapt for human cohort data. A regularization term in the covariance matrix with MPTGA (equation (3)) may lead to a better replication rate, lower false positive rate, while being less sensitive in detecting temporal eQTL (Figure S3a, S14a, b). The two Gaussian regression models (LR, Cubic) were more sensitive in detecting temporal eQTL but exhibited a lower replication rate and higher false positive rates compared to MPTGA (Figure S3a, S14a, b). LR and Cubic, without a regularization term, attempted to fit as much of the variance as possible, making them prone to overfitting ^3^ (Figure S15). The AR(1) approach, serving as the submodel for temporal eQTL detection in a tool called *TGCT* ^2^, is commonly used to detect temporal associations between two long time series, whereas our data consists of many short time series from multiple individuals^4^. The difference of dataset may explain why the AR(1) method performed least favorably.

### Supplemental Methods

#### Stationarity test on longitudinal observational data

To reduce the occurrence of the spurious correlation in subsequent temporal association analysis, we test the stationarity of longitudinal series data on the 17,236 gene-expression traits from multiple individuals based on the Augmented Dickey-Fuller (ADF) unit root test^5^. More exactly, ADF test on the longitudinal transcriptomic data was conducted under the formula and hypothesis below with the assumption of first order auto regression model, AR (1),

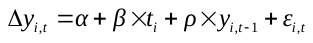

*H_0_*: *ρ* = 0

*H_1_*: *ρ* ≠ 0

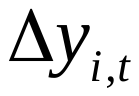
 denotes the first order difference in individual *i* at time *t* for a temporal trait *y*,
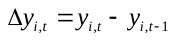
.

*α* denotes the constant intercept term.

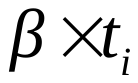
denotes the time trend term.

*ρ* is the coefficient of
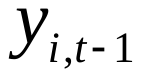
, representing the impact of
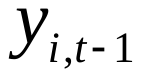
 on
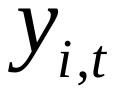
.

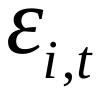
 denotes the error term, representing the random disturbance of the model.

If the null hypothesis *H_0_*: *ρ* = 0 is accepted, indicating that the time series has a unit root and is non-stationary. *H_0_*: *ρ* = 0 is tested based on *t*-statistic and *P* values is computed by comparing with the critical value computed by MacKinnon's unit root^6^ approximate asymptotic distribution functions (MacKinnon's). *P*<0.05 were considered as significantly indicating that the time series is stationary. In total, 17,222 stationary longitudinal gene-expression traits passed the ADF test with the *P* < 0.05.

#### PCs correction for expression data

To accounted for hidden technical confounders (such as batch effects) and biological confounders (such as inter-individual differences in cell type composition or transcriptome-wide variance in the gene expression data), we used the first 33 transcriptome-wide expression principal components (PCs) on the discovery cohort and 37 PCs on the replication cohort as the latent covariates for gene expression levels (Figure S4). The number of PCs was selected by a Buja and Eyuboglu (BE) algorithm in *R* package *PCAForQTL^7^* (*v*0.1.0*,* Figure S4), which showed the proof of principal component analysis (PCA) outperforms popular hidden variable inference methods (such as surrogate variable analysis (SVA), probabilistic estimation of expression residuals (PEER), and hidden covariates with prior (HCP)) for molecular QTL mapping.

#### Parameters estimation in MPTGA

To solve equation 1-4 in main text method, we first expressed
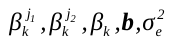
 and
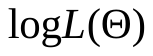
as functions of *ρ* as below, then looked for the critical point of
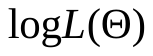
 which reached its maximum.

Notation (*Lin et.al. and Wu et.al*):

(1)
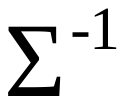
is a tridiagonal symmetric matrix. Its diagonal elements are
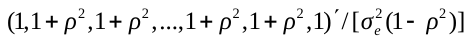
 and its second diagonal elements are all
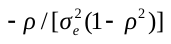
.

(2)
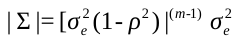
.

(3) Let
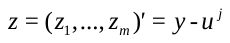
, then

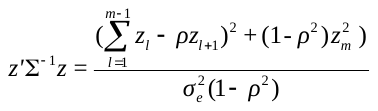

By taking derivative of
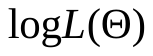
with respect to
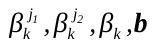
, the following linear system was obtained:

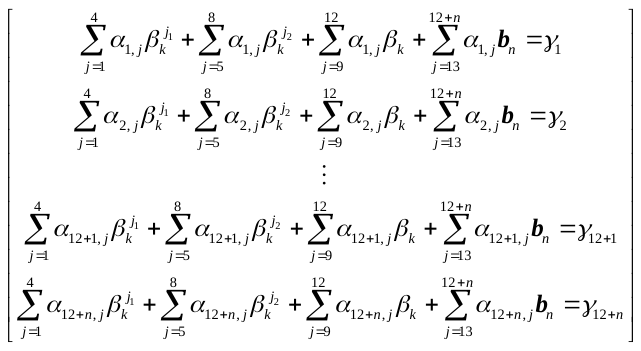

Let
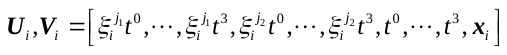
,

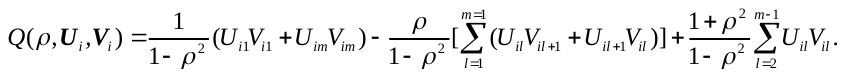

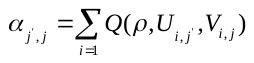
 and
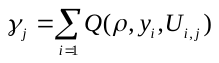
. Then, the coefficients for the linear system could be obtained by

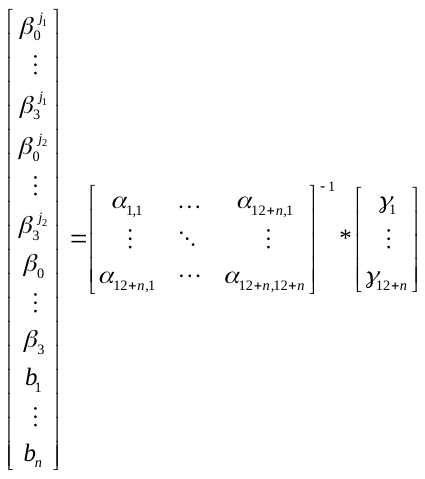

Taking derivative with respect to
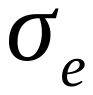
and let
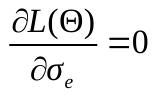
, then
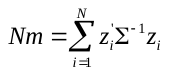
, *N* is the number of participants and m is the number of time points. We had
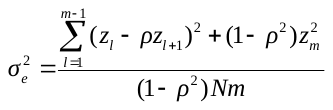
. Hence, we had
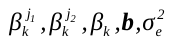
were expressed as a function of *ρ*. The log likelihood could be written as

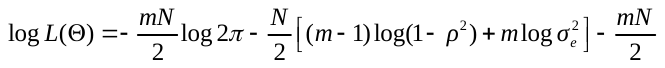
, thus, the MLE
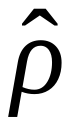
 could be obtained by looking for the critical point that maximizes
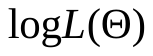
. Then the MLE for
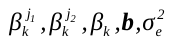
 could also be obtained.

#### Other method for temporal eQTL mapping

Union method. A straightforward approach described in Lin et al.^2^ which leverage gene expression data from the entire time series is to independently perform eQTL mapping at each time point. Then combining the results from analysis of all *m* time points at a locus as the following:
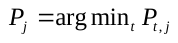
, where
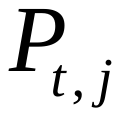
 is the *P* value for the gene expression trait *j* at the time point *t*. It means that if a gene expression trait was significantly linked to a locus at any of the *m* time points, the trait was linked to the locus in the union method.

Linear regression (LR) The LR model, a prevalent approach for time series data analysis, assumes independent variances at each time point (*ρ* = 0 in equation (3)), while MPTGA method assumes variances correlation stationarity (
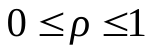
 in equation (3)). The model of LR for a temporal gene-expression trait is written as equation 1 with *K*=1. We performed an *F-test* to compare the reduced model (equation (5) with *K*=1) against the full model (equation (6) with *K*=1) to detect temporal eQTL association.

Cubic polynomial regression (Cubic) Francesconi et. al.^8^ leveraged cubic polynomial regression model to infer gene expression dynamics of *Caenorhabditis elegans* from single expression profiles in different genotypes. The variances of the cubic polynomial regression model at each time point are also independent (*ρ* = 0 in equation (3)) and can be written as equation (1) with *K*=3. Similar to LR, we compared the reduced model (equation (5) with *K*=3) against the full model (equation (6) with *K*=3) using *F-test* to detect temporal eQTL association.

First order auto regression, AR(1) The AR(1)^2^ model is another common approach for modeling time series data. To access whether gene-expression trait *y* is temporal associated with a genetic locus, we compared a null model

*H_0_*:
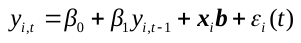

vs. a full model as

*H_1_*:
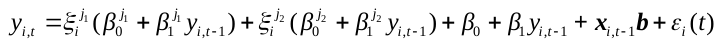
.

Where
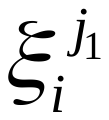
,
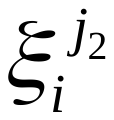
=0,0/0,1/1,0 is binary encoded genotype.
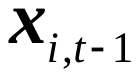
 is a (1×n)-design matrix of individual *i* for n covariates at time *t*-1, *b* is the vector for n covariate effects. We compared the reduced model *H_0_* against the full model *H_1_* using *F-test* to detect temporal eQTL association in AR(1).

#### Comparison of different genotype encoding forms

The genotype of SNP in human are commonly expressed as 0/1/2. Using this form of genotype data, temporal eQTL or dynamic eQTL can be detected by comparing between full model *H_1_* and null model *H_0_* or *H_0_’* as below:

*H_1_*:
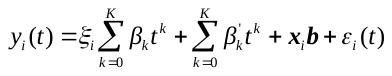

*H_0_*:
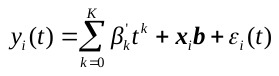

*H_0_’*:
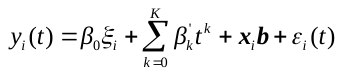

Where
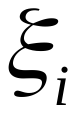
 is the genotype of individual *i*. The meaning of the other parameters is similar in equation (1) in Methods.

We identified and replicated 637,925 temporal *cis*-eQTLs and 3 dynamic *cis*-eQTLs when the genotypes were expressed as 0/1/2. when the genotypes were expressed as binary encoding, 00/01/10, 774,533 temporal *cis*-eQTLs and 6,660 dynamic *cis*-eQTLs were replicated. More parameters are needed for dynamic *cis*-eQTLs mapping when genotypes expressed as 00/01/10 than 0/1/2, which may be the reason of binary encoding genotypes identified more dynamic *cis*-eQTLs.

#### Simulation pipeline for evaluation of temporal and dynamic eQTL mapping approaches.

We conducted simulations of temporal gene-expression traits using a multivariate normal distribution. The mean vector was predicted by
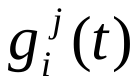
 with and without temporal genetic terms in the equation (1) to mimic the situation of existence and absence of temporal eQTL effects. The temporal eQTL effect of the simulation traits were randomly selected from a pool of 171,390 temporal *cis*-eQTLs, all of which were found to be significant in all five temporal eQTL mapping approaches. The coefficients in
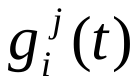
 was derived based on the Cubic approach. Additionally, we simulated 10 covariates through random sampling from the distribution *N*(1,0.5), while the covariate coefficient vector *b* were randomly sampled from *N*(0,0.5). Genotype were simulated from binomial distribution *B*(2, MAF), with MAF representing the minor allele frequency of the selected temporal eQTL SNPs. The covariance matrix was modeled as above in equation (3), where *ρ* was simulated with a distribution
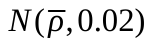
, with
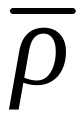
 ranging from 0.1 to 0.9 at 0.1 intervals. Subsequently, we simulated a total of 18,000 datasets, each containing a five-point time series and 300 participants, with and without temporal eQTL effects. In addition, we simulated temporal gene-expression traits to represent scenarios with and without dynamic eQTL effects, allowing for a comparison of the performance of dynamic eQTL mapping using MPTGA, LR, and Cubic, following a similar simulated pipeline as described above. True and false positive rate, as well as the area under the curve (AUC) were leveraged to evaluate the accuracy of temporal eQTL and dynamic eQTL detection. Comparison between AUCs was conducted by *DeLong’s* test^9^ in R package *pROC* (v1.18.0). The *P* < 0.05 is considered as significance.

#### Evaluating temporal QTL mapping methods

In order to compare the performance of various approaches in detecting temporal genetic associations in a time course human longitudinal study, we utilized the five temporal QTL mapping approaches to a set of simulated data. To simulate the interdependence of time points in a time series, we generated time-series data with varying strengths of correlation among the residuals at each time point (auto-correlation), ranging from 0.1 to 0.9 with an interval of 0.1. Each interval consisted of 1,000 patterns, resulting in a total of 18,000 simulated time-series patterns. These patterns were designed to mimic nonlinear time series data with multiple confounders and included both patterns with and without temporal eQTL effects observed by the Cubic approach in the discovery dataset.

The results of the temporal genetic association analysis indicate that the Union, LR, and Cubic approaches exhibited higher true positive rates than the MPTGA (Figure S15a), while the AR(1) performed the least favorably. Conversely, the MPTGA and AR(1) demonstrated lower and consistent false positive rates compared to the other approaches as the auto-correlation strength varied (Figure S14b). Specifically, the false positive rates of the LR and Cubic approaches increased with stronger auto-correlation (Figure S14b). Overall performance evaluations (Figure S14c, Additional file 7: Table S6) revealed that MPTGA achieved the highest AUC value and outperformed the LR, Cubic and AR methods (*P* < 0.01, Methods) in scenarios involving strongly auto-correlated data (average auto-correlation proportion = 0.9).

#### eQTL SNPs annotation

We used *snpEff* (v5.2, https://pcingola.github.io/SnpEff) for our findings of dynamic and additive *cis*-eQTL SNP annotation and variant effect prediction with the command

*java -jar ./snpEff.jar -v GRCh38.86.*

See Table S7,S8 for the variant annotation and effect prediction of each eQTL SNP.

#### Instrument variable selection

We excluded temporal eQTL SNPs that were associated (*P* < 0.05) with candidate confounders of neurodegenerative-related phenotypes (Such as age, sex and et.al.) in the discovery dataset. *P* value was calculated by testing the significance of correlation between a temporal eQTL SNP genotype data and a confounder using linear or logistic regression model. Linear and logistic model were conducted in python package *statsmodels* (v0.14.0).

#### Cointegration test

We performed unit root test^5^, which was described in the stationarity test in Methods, on the residual *ε* of the significant causality VAR models in equation (10) to test the cointegration^6^ between two lagged correlation temporal traits.

#### Optimal lag selection

For a causal gene which was significant in VAR models with distinct lags, we used Schwarz criterion^10^ (SC) for selecting optimal lag as the optimal temporal eQTL SNP-gene-outcome causality model.

SC value can be calculated as

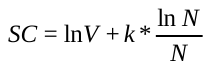

Where *V* is the estimated white noise variance; *k* is the order or lag; *N* is the number of observations.

#### Power estimation in the causality analysis of 2sGen-GPS

The temporal causal effect size of a gene in 2sGen-GPS is quantified by F-value. Hence, we used function *FTestPower* in the python package *statsmodels* (v0.14.0) to estimate the power of temporal causal effect size in our findings.

#### Method of DBM and microstructural gradients features measurement

The neuroanatomic features utilized in this study were obtained through the application of deformation-based morphometry (DBM)^11^. DBM relies on nonlinear and intensity-based registration procedures that spatially normalize the entire brain to a standard template. DBM does not make assumptions about the distributions of gray matter or white matter and retains the complete MRI data. An important advantage of the DBM method is its ability to identify subcortical neuroanatomic features^12^, Previous research has demonstrated that DBM can effectively detect structural tissue alterations in individuals with early-stage PD^13^.

T1-weighted MRI scan acquisition parameters are detailed elsewhere (www.ppmi-info.org). The T1-weighted MRI images were preprocessed with the Computational Anatomy Toolbox (CAT 12) (dbm.neuro.uni-jena.de/cat12/), which is an extension of SPM12 to provide computational anatomy. All images were corrected for bias, noise, and intensity and linearly and then nonlinearly registered to the Montreal Neurological Institute 152-2009c template. Then the determinant of the jacobian transformation matrix maps were calculated to estimate the local volume in each voxel (DBM values). Finally, the obtained preprocessed volume-based DBM data from 171 PD cases with 559 visits were resliced to the aal3^14^ template using SPM12 for brain region labeling.

We used *mrGrad* (https://github.com/MezerLab/mrGrad) for calculating the microstructural gradients features of specific brain region. The *mrGrad* was an automatic procedure^15^ and have been leveraged to generate qMRI functions along the main axes of a subcortical structure at the single-subject level. The calculating procedure in *mrGrad* is as following.

Automatic axis computation

Given an ROI mask (aal3 in this study), the algorithm computes the SVD of the ROI’s voxel 3D image coordinates to find the main three orthogonal axes (i.e., the eigenvectors) of the structure. The SVD algorithm solves for *M = U Σ V **

where *M* is an m × 3 matrix of the ROI’s *m* centered image coordinates, *Σ* is a diagonal matrix with the singular values of *M*, and *U* and *V* are matrices whose columns are the left and right singular vectors of *M*, respectively (or the orthonormal eigenvectors of *MM** and *M*M*, respectively). Therefore, the columns of the 3 × 3 matrix *V* define the main orthogonal axes of the ROI, based on its anatomical shape.

Axis segmentation

For each of the three main axes, the ROI is segmented with equal spacing. For this purpose, the data edges are defined as the two hyperplanes defined by the two extreme data points of the data with respect to the axis and by the axis as a normal to the plane. The data are then segmented by *n* − 1 parallel hyperplanes equally spaced between the two data edges. Voxels are then classified to *n* segments based on criteria of distance from planes. In our analysis, we chose the default *n* as 7.

Two examples of microstructural gradients features (left caudate nucleus 7 and left substantia nigra pars reticulata 3 in PC1) are showed in Figure 3f.

#### Study population and rs11241912 genotyping

Whole-genome sequencing data were obtained from the Accelerating Medicines Partnership PD (AMP-PD) project (https://app.terra.bio/#workspaces/amp-pd-public/AMP-PD-In-Terra) ^16^, comprising 100 post-mortem brain donors collected from four brain banks: Mount Sinai Brain Bank, University of Miami Brain Endowment Bank, Harvard Brain Tissue Resource Center, and Udall Center of Excellence for PD Research. There was a similar distribution of sex and age for each brain bank. The cohort comprised 75 PD donors stratified using Braak PD staging, which tracks the spread of Lewy body pathology, and 25 neuropathologically confirmed controls. Genomic DNA was extracted from the primary visual cortex (PVC) for whole-genome sequencing.

#### Single-nucleus RNA sequencing (snRNA-seq) data preprocessing and quality control (QC)

The snRNA-seq data processing, clustering, and cell type annotation were performed using *Scanpy* (V1.10.4) (https://github.com/scverse/scanpy) ^17^ through a four-step QC pipeline. The first QC stage implemented stringent cell-level filtering criteria: (1) unique molecular identifier counts (1500–110 000), (2) gene counts (1100–12 500 genes), and (3) mitochondrial gene percentage (<2%). Ambient RNA contamination was assessed by monitoring non-coding RNA signatures, including ribosomal RNA, small non-coding RNA, pseudogenes, and the long non-coding RNA *MALAT1*. Potential doublets were identified and removed using *Scrublet* (v.0.2.3) (https://github.com/swolock/scrublet) with default parameters ^18^. The second QC stage filtered lowly expressed genes (detection rate < 0.05% across nuclei). The third QC step excluded cell types with <50 cells to ensure robust downstream analysis. The final QC step removed mismatched cells based on a comparison of cell annotation results from the reference and our annotation results. Finally, a dataset was obtained containing 42 samples from the prefrontal cortex (PFC) and 230,806 cells. Gene counts were normalized to the total counts per cell, multiplied by 10 000, and then log-transformed. Highly variable genes were identified using specific filtering parameters (min_mean = 0.0125, max_mean = 3, min_disp = 0.5), and these genes were used to compute low-dimensional embeddings via Uniform Manifold Approximation and Projection (default parameters using 50 principal components and 15 nearest neighbors). Clustering was performed using the Leiden algorithm (resolution = 0.04, 10 iterations), resulting in preliminary cell clusters.

#### scRNA-seq cluster marker analysis and cell-type annotation

Cell-type annotation was conducted using *CellTypist* (V1.6.1) (https://github.com/Teichlab/celltypist)^19^, a logistic regression classifier optimized with the stochastic gradient descent algorithm. First, we trained the model using an snRNA-seq dataset from the PFC of the brain, annotated as part of the ROSMAP project (<http://compbio.mit.edu/ad_aging_brain/>) ^20^. The same QC procedures applied to our dataset were also implemented for the ROSMAP dataset before training using the *celltypist.train* function. Subsequently, the trained model was used to predict cell types and subtypes, based on the *celltypist.annotate* function with the parameter “majority_voting = True” to provide consistent labels.

#### Region- and cell-type-specific differential expression analysis between rs11241912 carrier and non-carrier

Differential expression (DE) analysis between rs11241912 carriers and non-carriers was performed using the *Scanpy* framework. Raw counts were normalized by total library size per cell (*scanpy.pp.normalize_total*) and log1p-transformed (*scanpy.pp.log1p*). To identify cell-type-specific DEGs across the five brain regions (DMNX, GPI, PMC, PFC, and PVC), we utilized the *scanpy.tl.rank_genes_groups* function, employing a two-sided Wilcoxon rank-sum test. Analysis was stratified by seven major cell types: Ast, Exc, Mic, Inh, Oli, OPC and End. Significant DEGs were defined as those with a Benjamini-Hochberg adjusted P-value (FDR) < 0.05 and an absolute *log_2_* fold change (*|log_2_FC|* > 1).

#### ROSMAP cohort quality control and genotype imputation.

Genotype data for ROSMAP were obtained from 1,708 subjects of European ancestry using the Affymetrix GeneChip 6.0 (reference genome: hg18), comprising 750,153 SNPs (https://www.synapse.org/#!Synapse:syn3157325). We conducted subject and SNPs quality control using PLINK (v1.90 beta) (PMID: 17701901). The quality control process involved several steps: excluding 27,984 SNPs not present in the hg38 assembly after Liftover conversion (https://genome.ucsc.edu/cgi-bin/hgLiftOver), as well as excluding 86,697 SNPs and 24 subjects with an overall missingness > 0.05. No subjects were excluded due to relatedness (PI_HAT > 0.1875) or gender mismatch. Additionally, 146 SNPs with Hardy-Weinberg equilibrium (*P* < 10^-6^) and 67 SNPs with mishap (*P* < 10-9) were excluded, along with 16 subjects with a heterozygosity rate (F value) > 4 s.d. from the mean. Furthermore, no population outliers were detected through principal component analysis (PCA). Ultimately, 1,668 subjects and 635,657 SNPs with MAF ≥ 0.001 remained for genotype imputation. Genotype imputation was carried out using Eagle2 (v2.4) and Minimac4 (v1.0.0) on the TOPMed Imputation Server (PMID: 27571263), with the TOPMed panel (version R2) selected as the reference panel. This reference panel comprises 194,512 haplotypes and 308,107,085 autosomal SNPs. Details of the TOPMed Imputation Server pipeline can be found at https://imputation.biodatacatalyst.nhlbi.nih.gov. Following genotype imputation, 10,097,225 imputed variants with R^2^ ≥ 0.3 and MAF ≥ 0.001 were retained for further analysis.

### Supplemental Figures

#### Figure S1. Schematic of the 2sGen-GPS framework

The 2sGen-GPS framework integrates longitudinal multi-omics through a two-stage analytical pipeline. Stage I: Characterization of dynamic genetic regulation. Using the MPTGA (Multivariate Polynomial Temporal Genetic Association) module, we characterized the temporal architecture of *cis*-eQTLs across two longitudinal blood transcriptome datasets. This stage involved distinguishing between constant (additive) and nonlinear dynamic regulatory patterns, capturing how genetic effects on gene expression evolve over time in both healthy individuals and PD patients. Stage II: Inference of temporal genetic causality. To bridge molecular dynamics with clinical progression, we utilized significant temporal cis-eQTL SNPs as instrumental variables (IVs). These IVs enabled the longitudinal imputation of individual-level gene expression trajectories, representing the genetically determined component of the transcriptome. Finally, we employed a vector autoregression (VAR)-based Granger causality model to estimate the temporal causal effects of these imputed expression trajectories on longitudinal disease phenotypes (e.g., MDS-UPDRS III and cognitive scores) and other omics (e.g., Proteomics and radiomics).
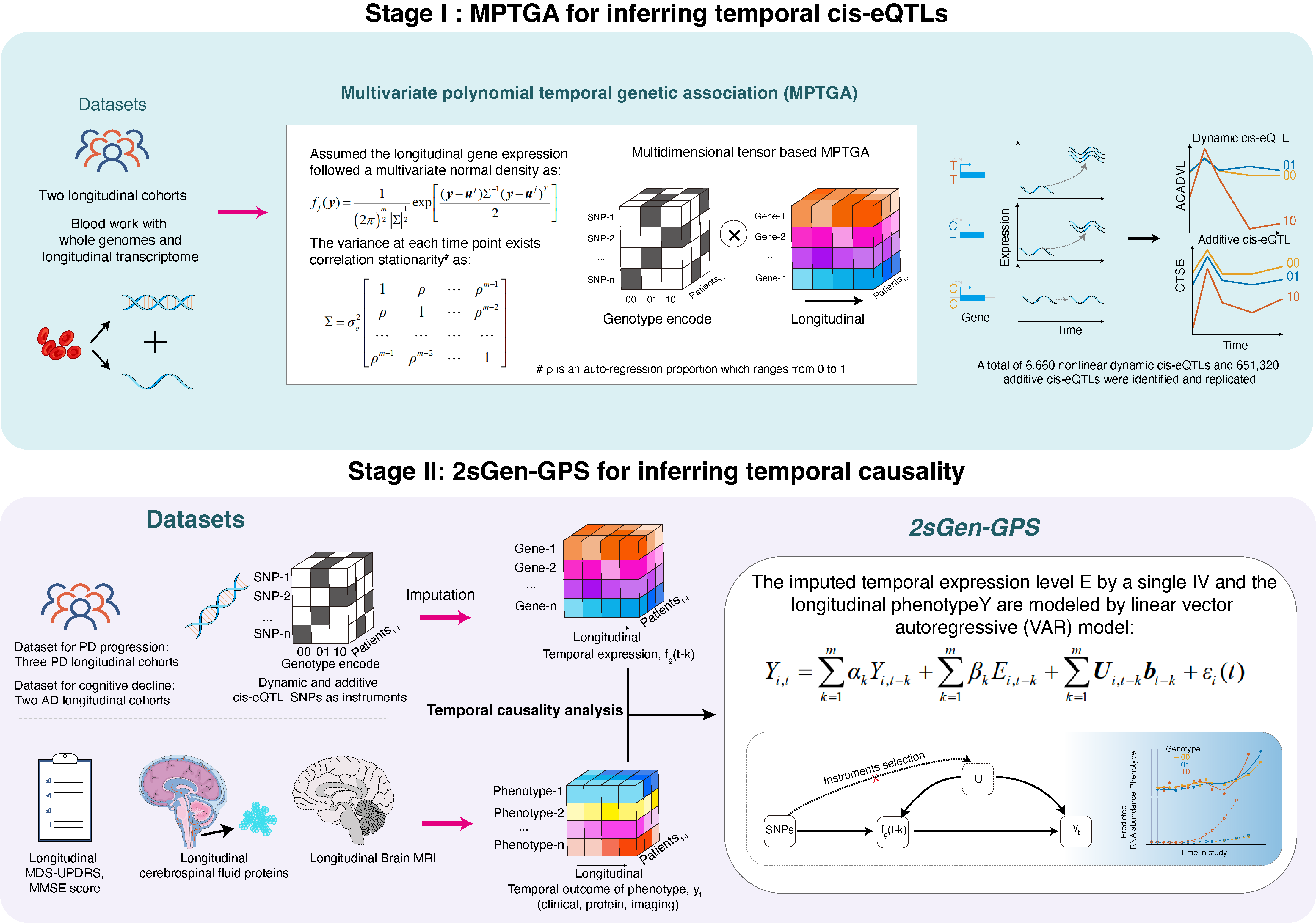

#### Figure S2. Overview of the datasets for temporal local (*cis*) eQTL and dynamic eQTL mapping.

a-c, Sex, age at baseline, diagnosis and race distribution in the discovery dataset and d-f in the replication dataset.

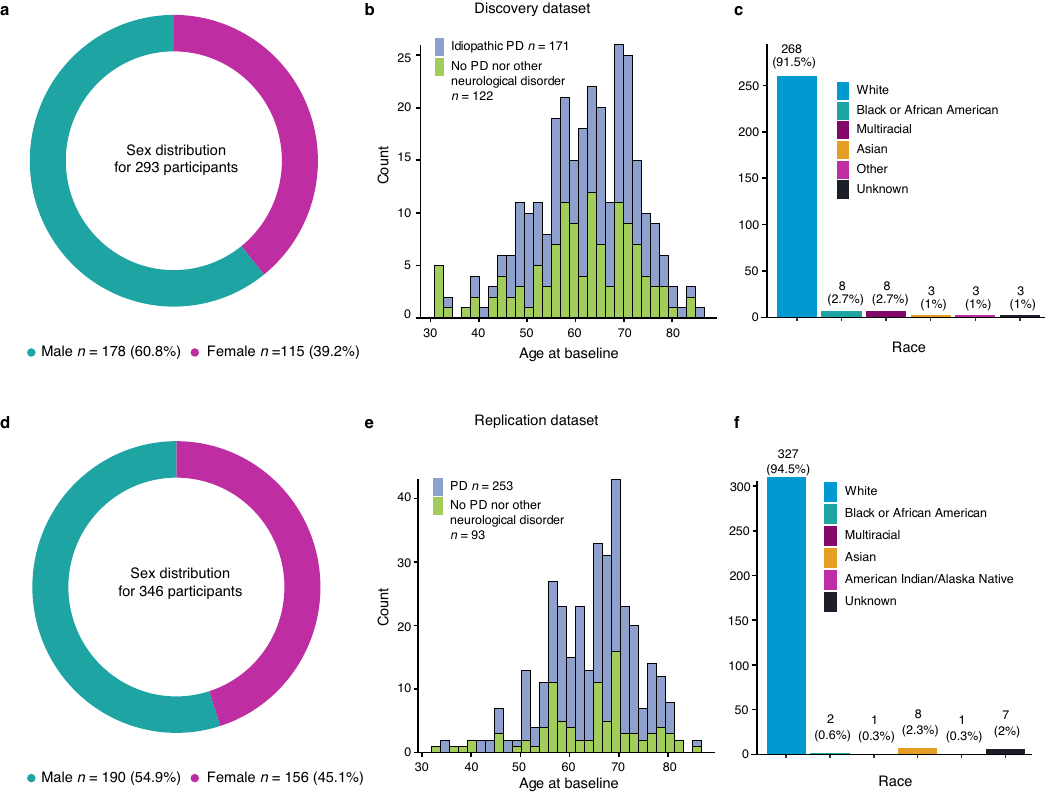

#### Figure S3. Results of temporal *cis*-eQTLs from five temporal eQTL mapping approaches in two blood longitudinal gene expression dataset.

a, Replication rate of the temporal *cis*-eQTLs identified by the Union, linear regression (LR), cubic polynomial regression (Cubic), first order auto regression (AR(1)) and MPTGA using independent replication dataset. Values above the percent of replication rate within each bar indicate the number of temporal *cis*-eQTLs in PPMI that have been validated in PDBP. b,c, The overlapped number of validated temporal *cis*-eQTLs and genes with temporal *cis*-eQTL in the Union, LR, Cubic and MPTGA approaches.

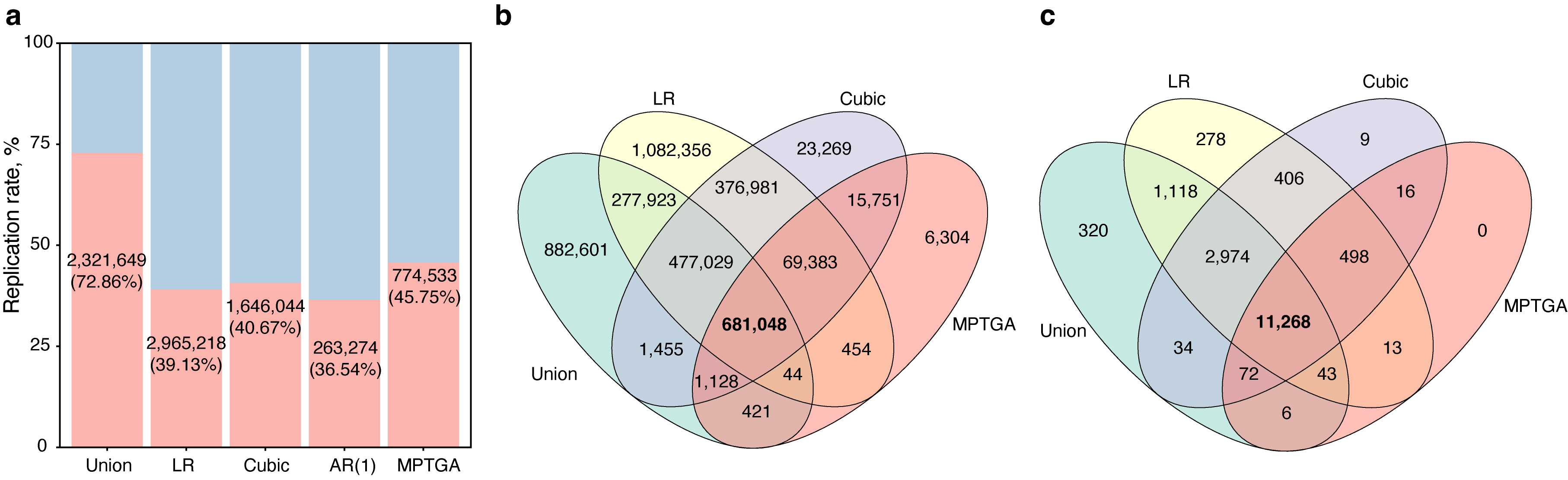

#### Figure S4. The choice of *K* latent factors to account for hidden batch effects and other technical and biological sources of transcriptome-wide variance in the discovery and replication dataset.

a, c, Proportion of variance explained (PVE) by principal components (PCs) in the discovery and replication datasets. The dots represent the PVE by a specific number of PCs; green vertical line indicates the choice of *K* latent factors using Elbow method and purple vertical line using Buja and Eyuboglu (BE) method^7,21^. b, d, The PVE of known covariates by *K* latent factors determined by BE method in the discovery and replication datasets.

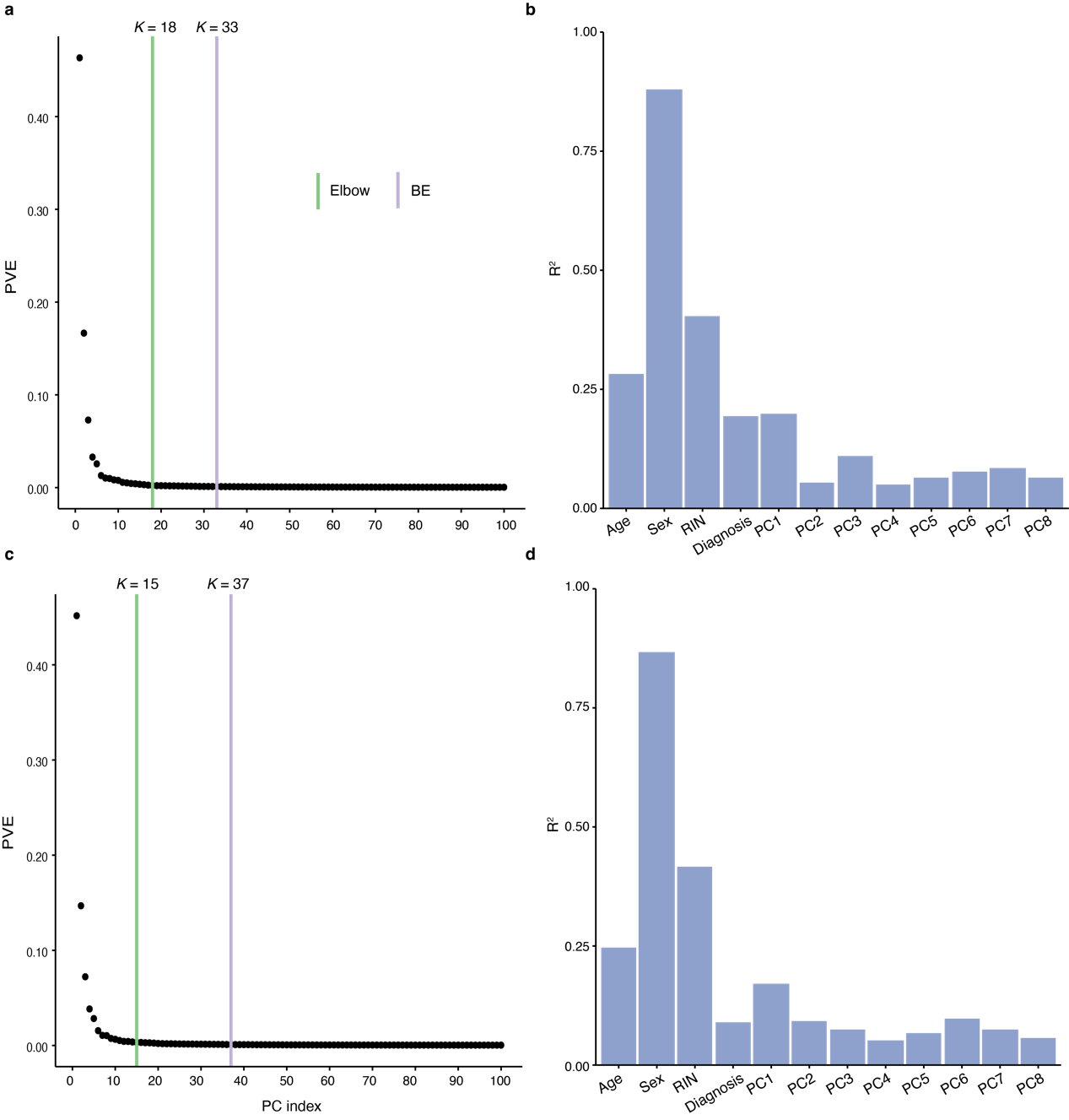

#### Figure S5. Example of nonlinear dynamic and additive *cis*-eQTLs.

a, Nonlinear dynamic *cis*-eQTL for three genes. b, Additive *cis*-eQTL for three genes. Colors indicate SNP genotype, with orange as the minor allele; the lines represent the mean of expression level trajectory during the study in the discovery dataset.

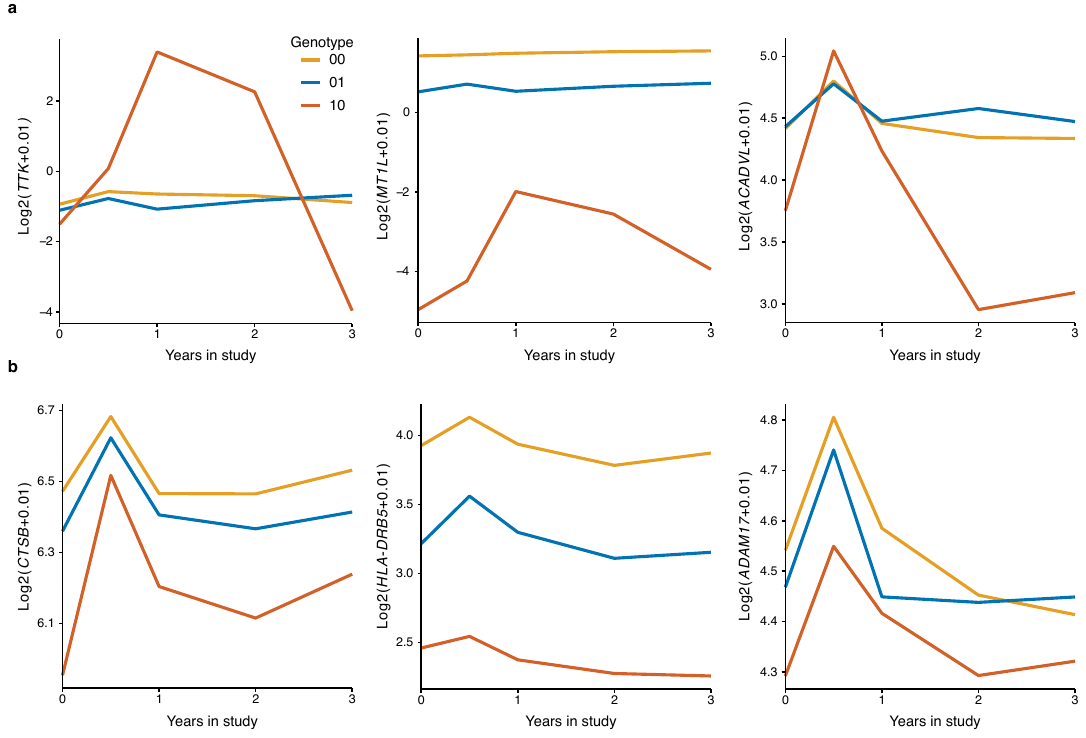

#### Figure S6. Power test for the 5,000 temporal *cis*-eQTL.

a, The examples of temporal *cis*-eQTLs with power = 80% (left) and 90% (right) using bootstrap approach. Density curve indicates the distribution of alternate hypothesis (H1); vertical solid line is the statistical significance cutoff, *α,* with *α*=0.001 (FDR < 0.05) in the discovery dataset and 0.05 in the replication dataset; area in the right side of vertical solid line under the density curve indicates the power of the temporal eQTL. b,c Number of temporal *cis*-eQTL with discovered and replicated power more than 80%. d, Evaluated the percent of temporal eQTL with enough power (power ≥ 80%) across different sample sizes.

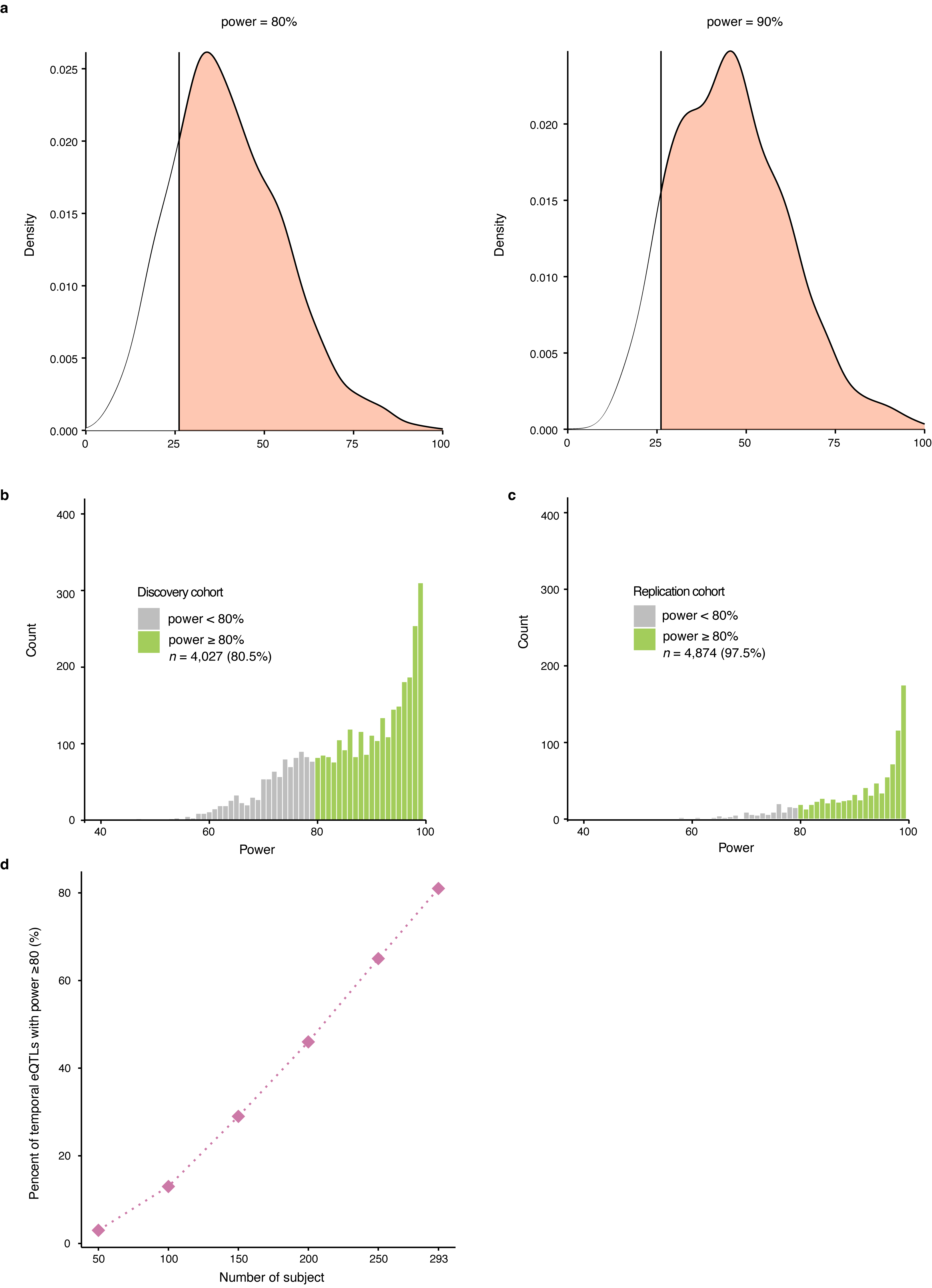

#### Figure S7. Evaluation of the temporal *cis*-eQTLs detected by MPTGA in the longitudinal blood RNA-seq.

a, The percentage of our whole blood *cis*-eQTLs overlapped with GTEx. Values above the percent of replication rate within each bar indicate the number of *cis*-eQTLs identified in both GTEx and our temporal eQTL mapping analysis. b, High correlation of PVE by temporal *cis*-eQTLs between discovery and replication datasets with a Pearson correlation coefficient = 0.97 (*P* < 0.001). Two-dimension (2d) histogram indicates the distribution of PVE by temporal *cis*-eQTLs in both datasets; histograms on the upper and right side of the 2d histogram showing the distribution of PVE by temporal *cis*-eQTLs in the discovery and replication datasets, respectively. Percentage of polymorphisms in the dynamic (c) and additive (d) *cis*-eQTLs in different regions of the genes.

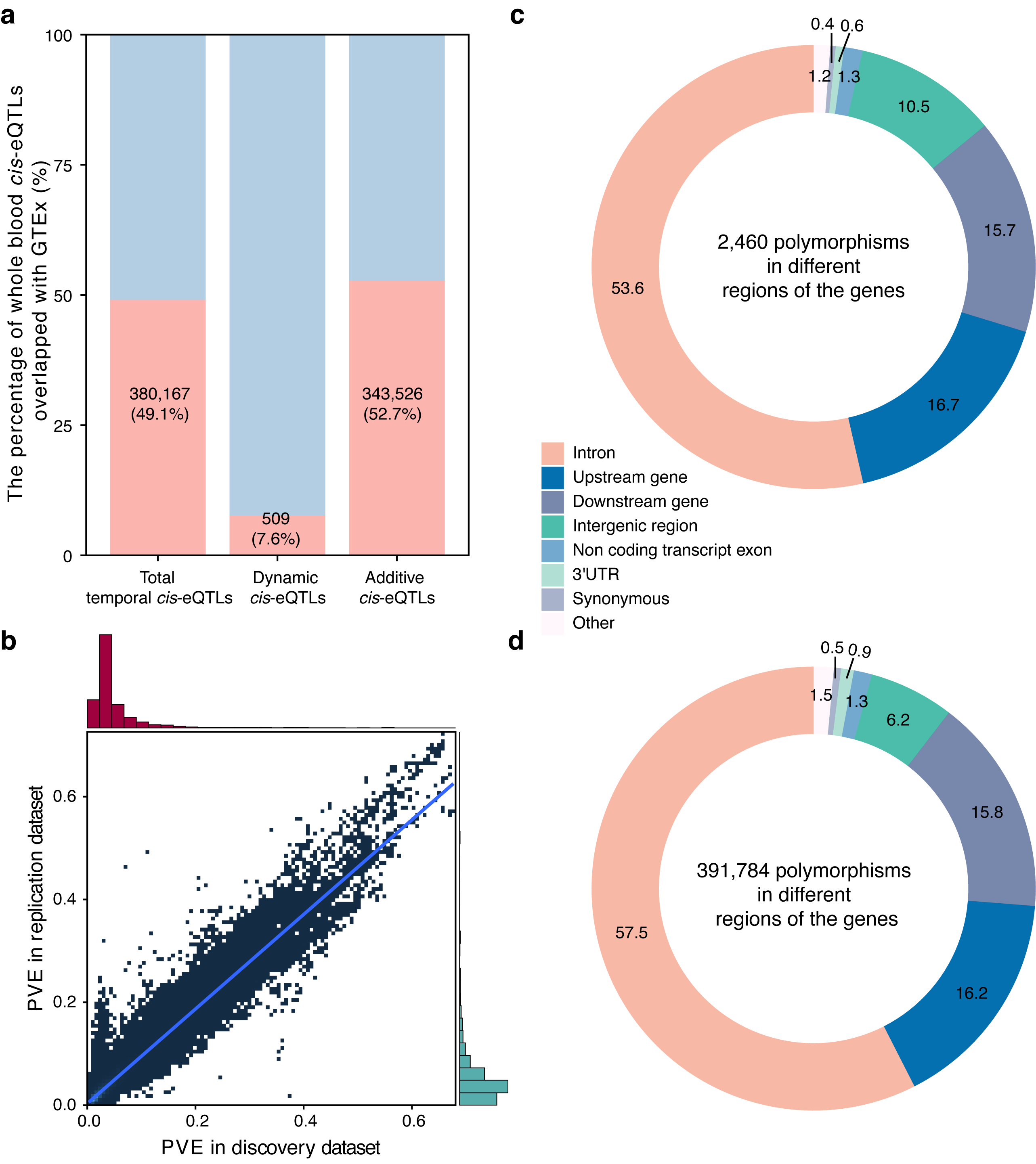

#### Figure S8. Overview of the datasets for temporal causality analysis in PD progression.

a, The distribution of longitudinal visits in different cohorts. b-d, The distribution of sex, age at baseline and race in different cohorts.

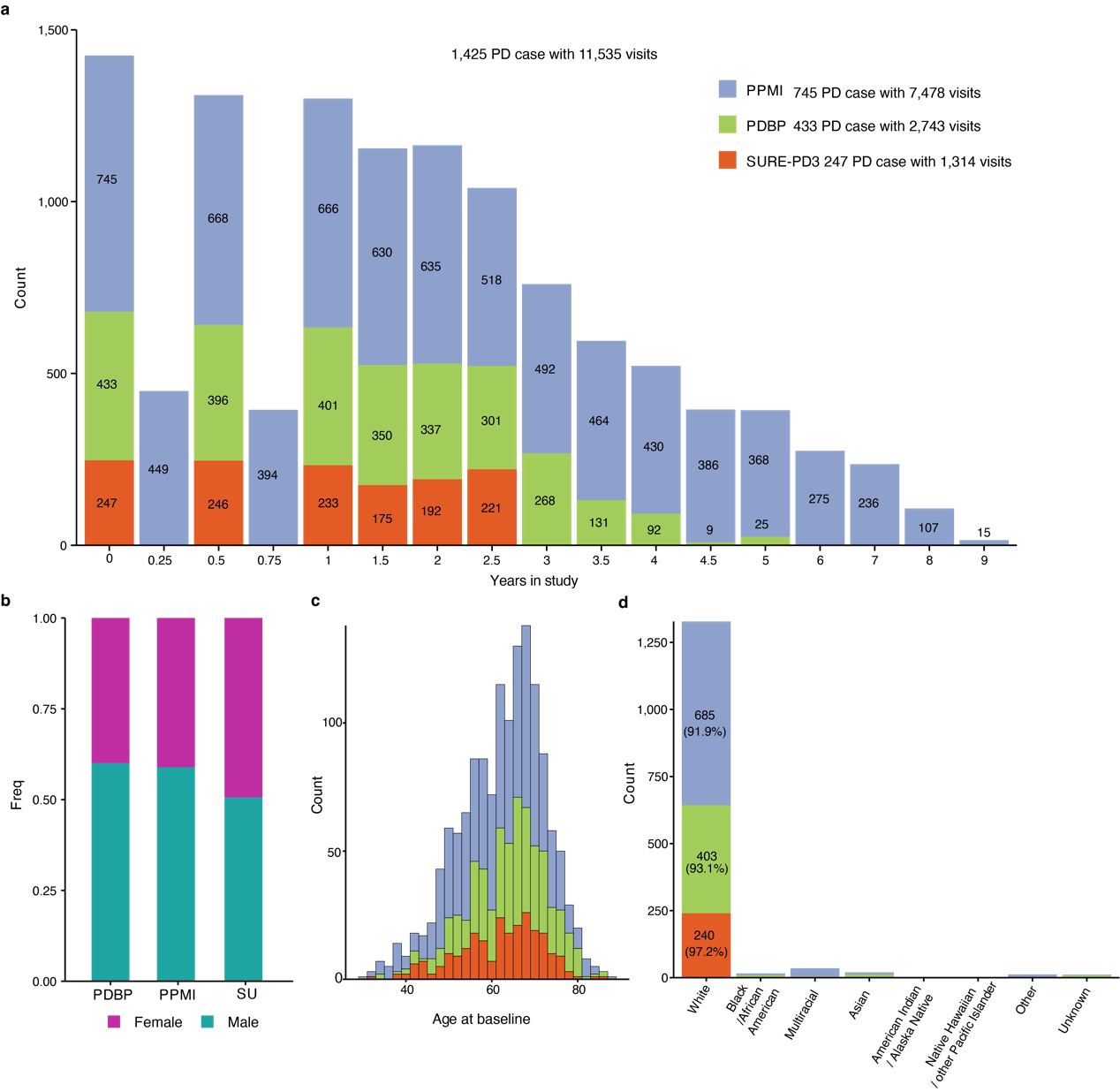

#### Figure S9. A workflow of temporal causality analysis in 2sGen-GPS.

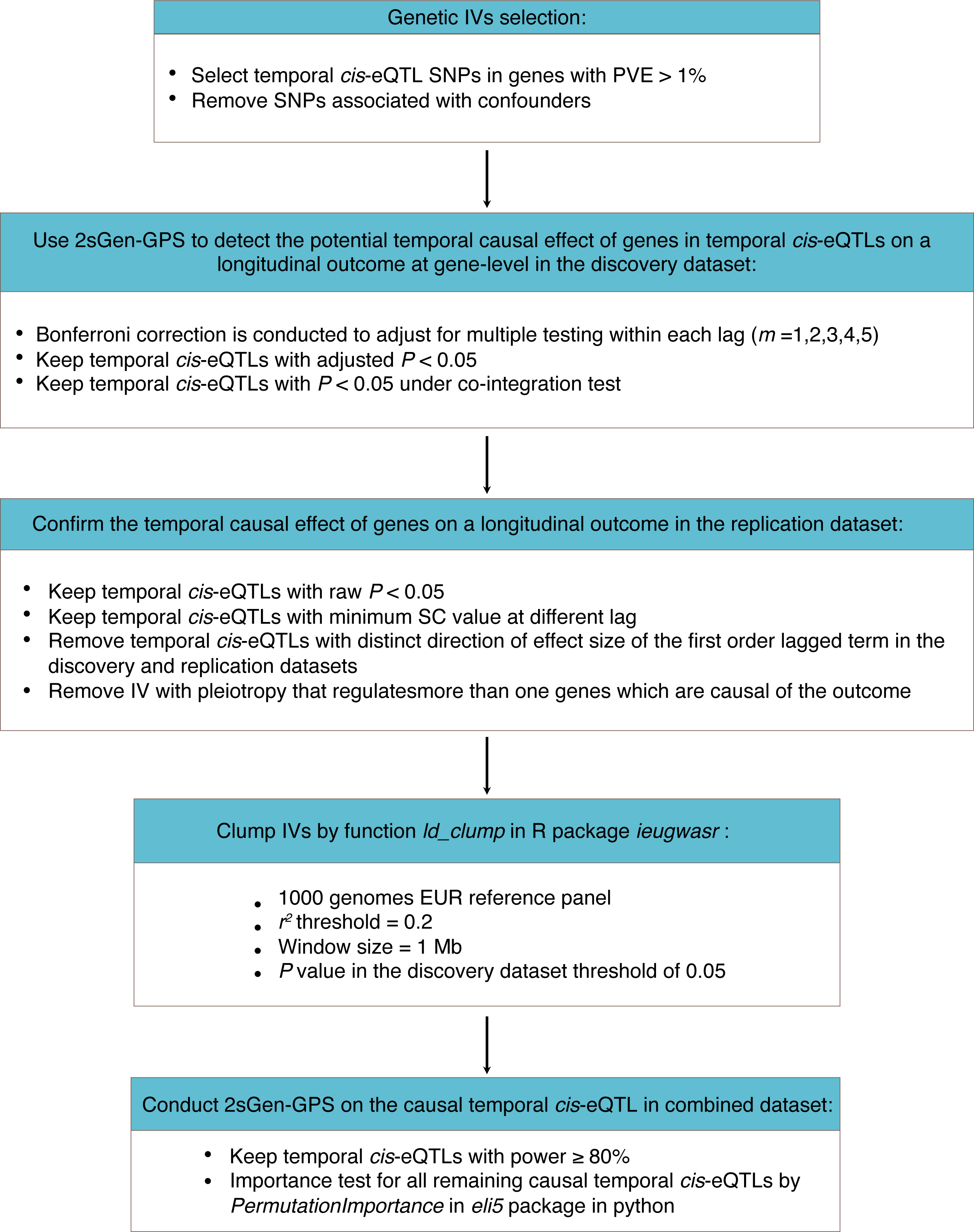

#### Figure S10. Temporal causality dynamic and additive *cis*-eQTL genes for motor impairment progression in the discovery (left) and replication (right) datasets.

a,b, Dynamic *cis*-eQTL genes (*AC051649.1* and *NR2C1*). c, additive *cis*-eQTL genes (*HLA-DRB5*). d, Gene Ontology (GO) enrichment analysis of the 427 blood causal genes of PD progression.

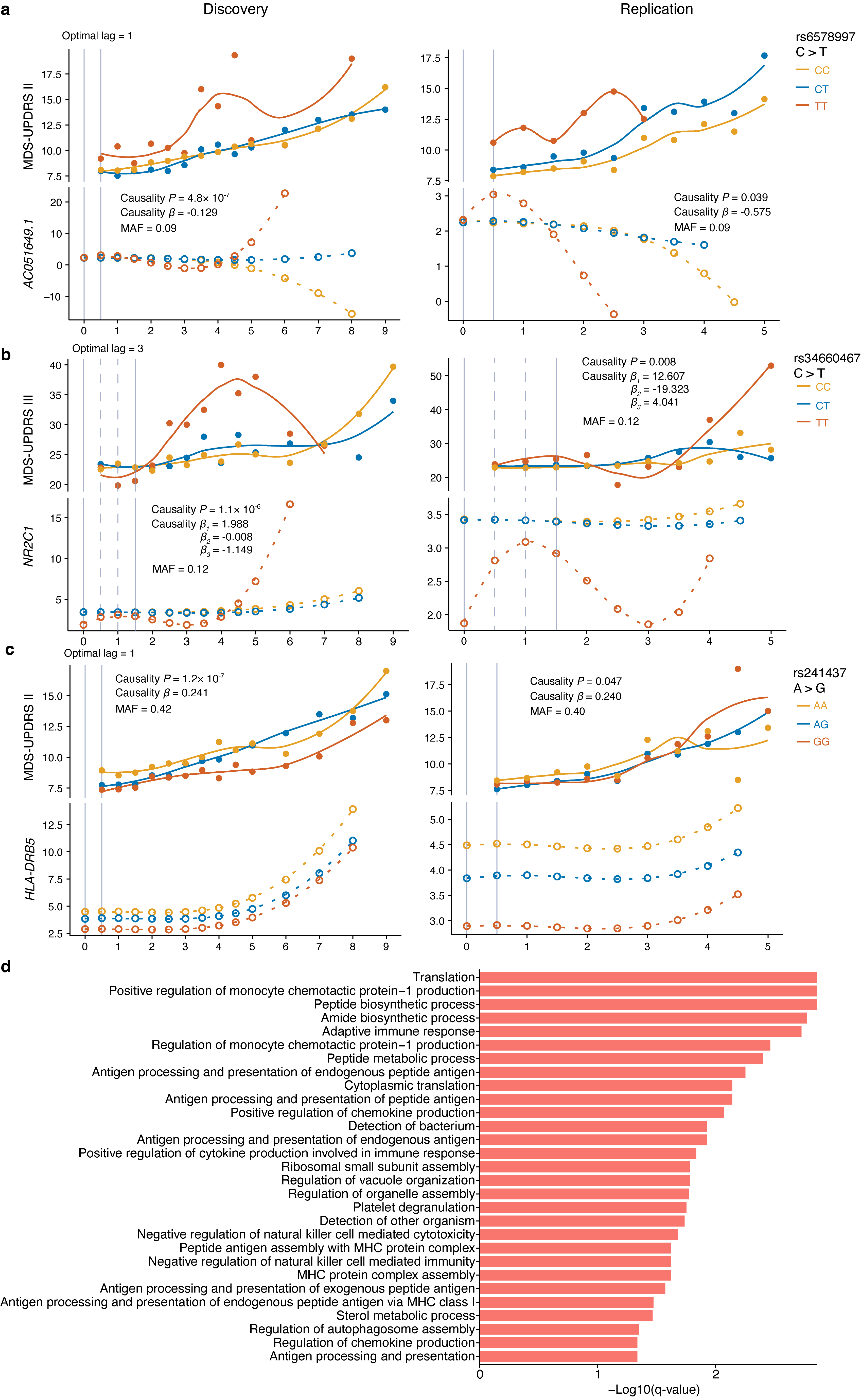

#### Figure S11. Shared temporal genetic effects between CSF proteins and PD motor symptoms in the discovery and replication datasets.

Visualization of gene *C5orf63* that is temporal causality with PD motor symptom (a) and CSF protein LRP1 (b) in the discovery (left) and replication (right) datasets during five years study. c, MDS-UPDRS III score trajectories over longitudinal visited time for PD patients harboring different rs11241912 genotypes (GG, GA, AA). Curves represent local polynomial regression (Loess) fits illustrating the progression of motor impairment. *N represents the number of subjects per genotype*.

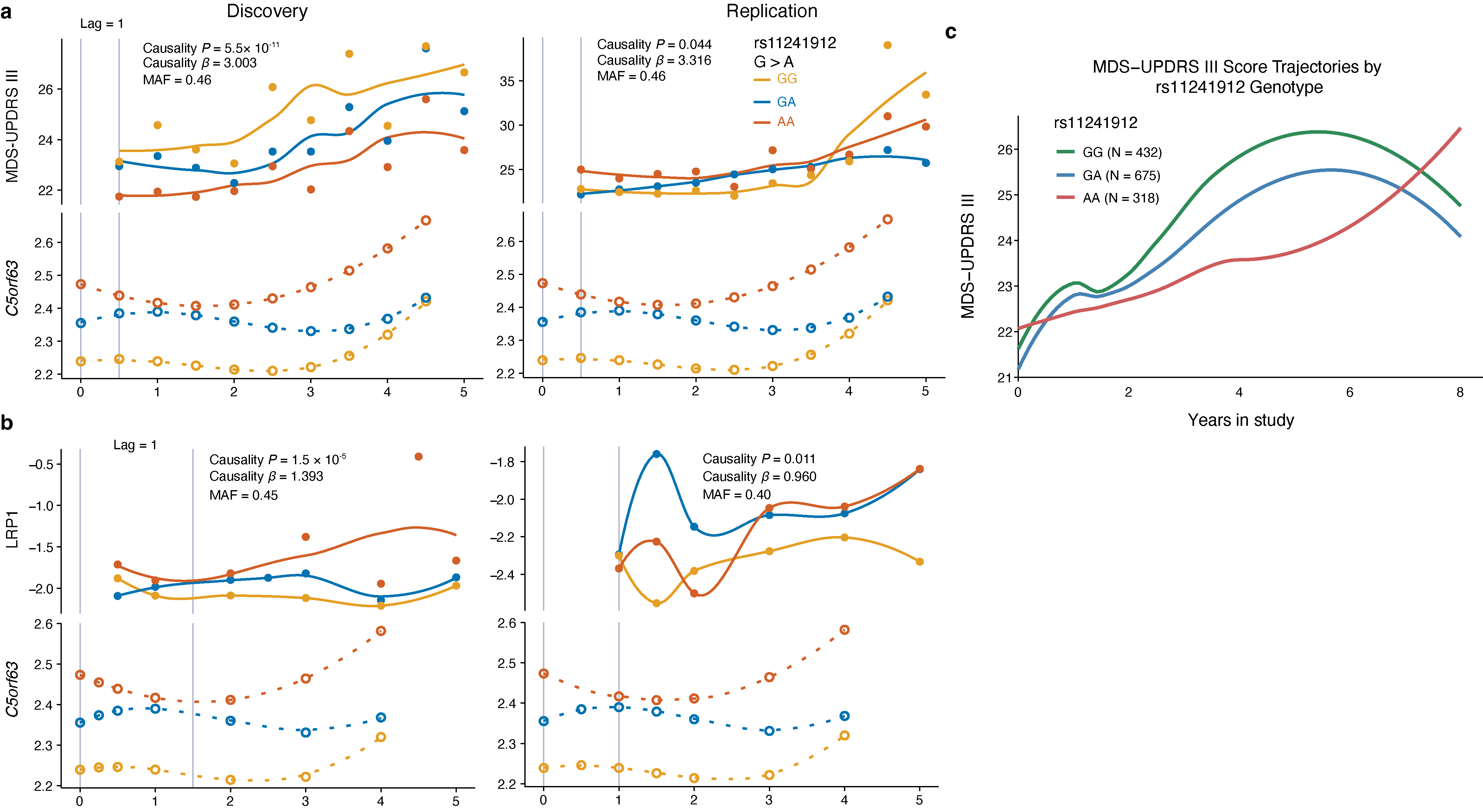

#### Figure S12. Overview of the datasets for temporal causality analysis in cognitive decline.

a, The distribution of longitudinal visits in both MAP and ADNI cohorts. b-d, The distribution of sex, age at baseline, and years of education in both MAP and ADNI cohorts.
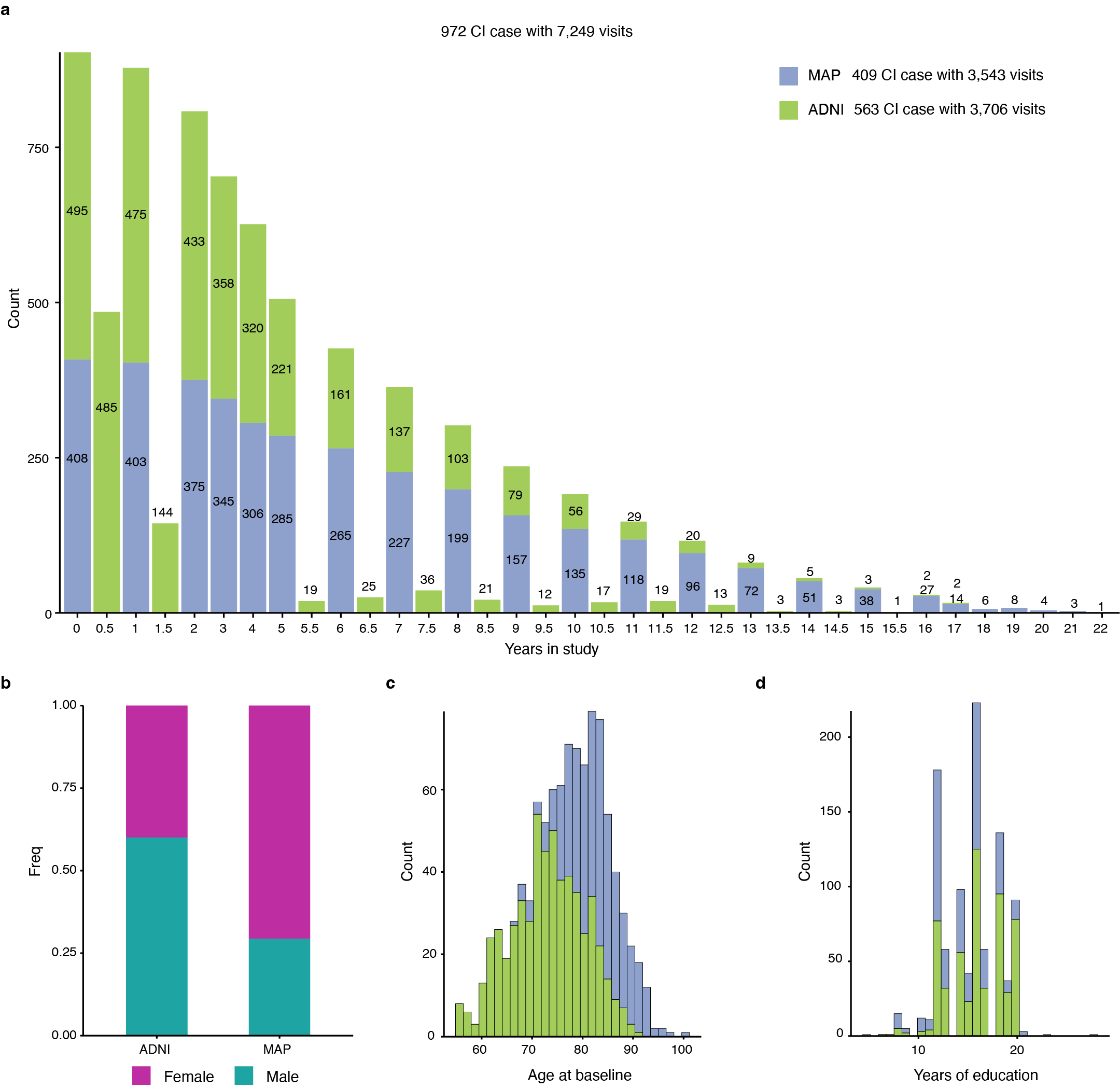

#### Figure S13. Temporal genetic causality analysis identifies dynamic driver genes for cognitive decline in the discovery and replication datasets.

a, Protein-protein interaction Gene Ontology (GO) enrichment analysis of the 381 blood causal genes of cognitive decline. Visualization of lagged correlation between dynamic *cis*-eQTL genes *SNCA* (b) and *RN7SL674P* (c) and mean of longitudinal MMSE linked to cognitive decline in the discovery (left) and replication (right) datasets.

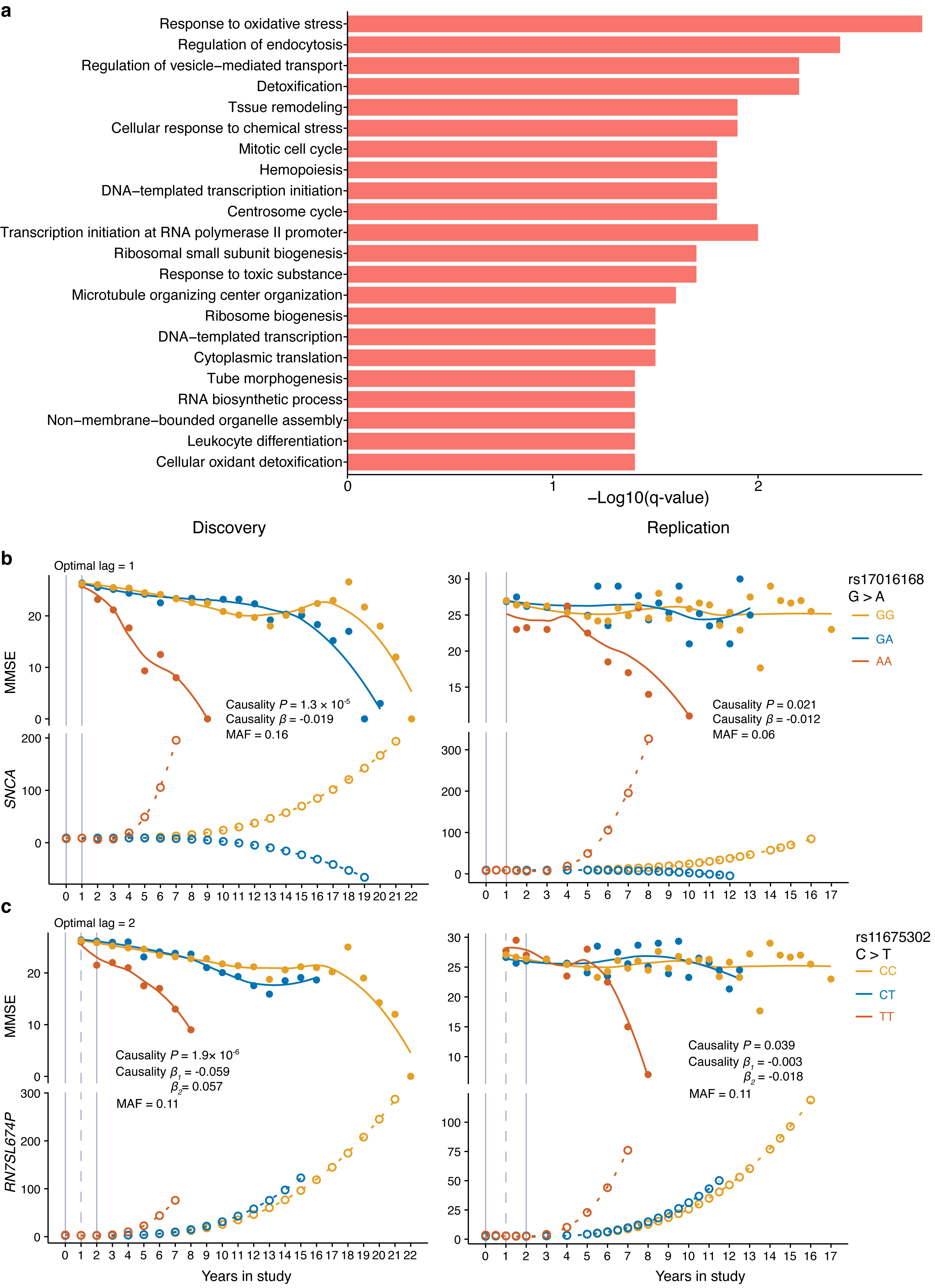

#### Figure S14. Comparison of temporal eQTL identification using five temporal eQTL mapping approaches in the simulated dataset.

a, b, True and false positive rate of the five approaches vary under different strength of auto-correlation proportion. c, Area under the curve (AUC) of the five approaches vary under different strength of auto-correlation proportion. All error bars indicate 95% CI.

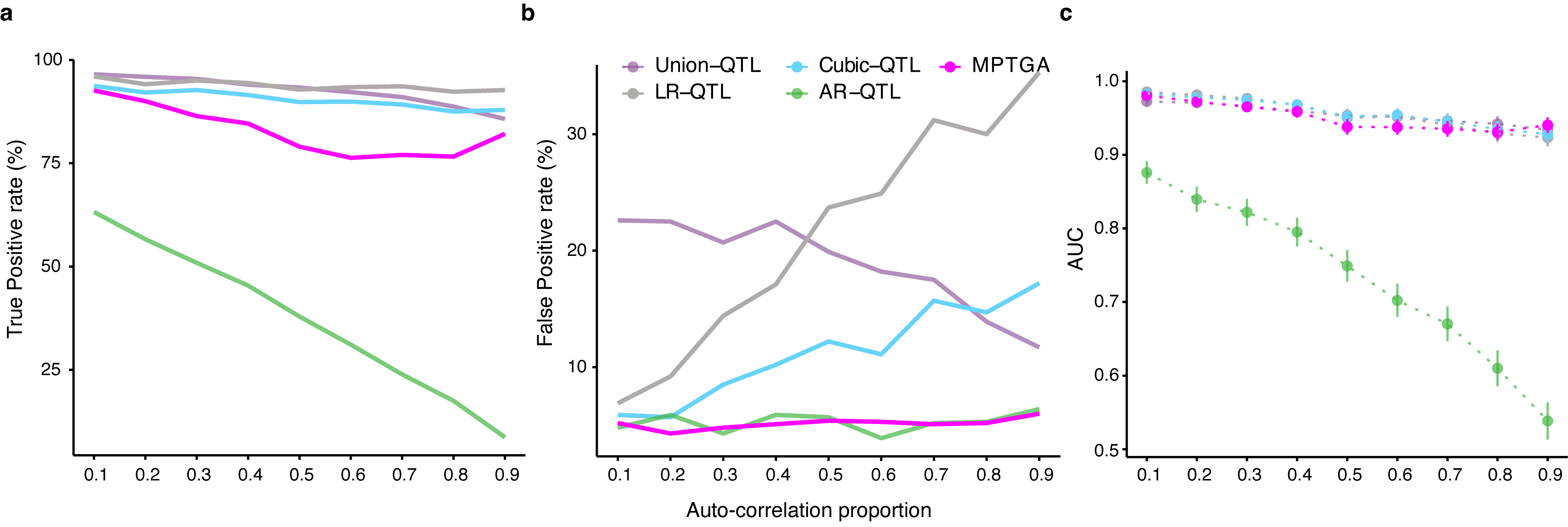

#### Figure S15. Comparison of goodness of fit on the models between MPTGA and Cubic approaches using *R^2^*.

Boxplot shows the *R^2^* of 171,390 temporal *cis*-eQTL models that are significant in MPTGA, Cubic and the other three approaches; two-sided *P* value from independent *t* test is shown. The upper and lower ends of the boxes represent the IQR of *R^2^*. The lines in the boxes represent the median *R^2^*.

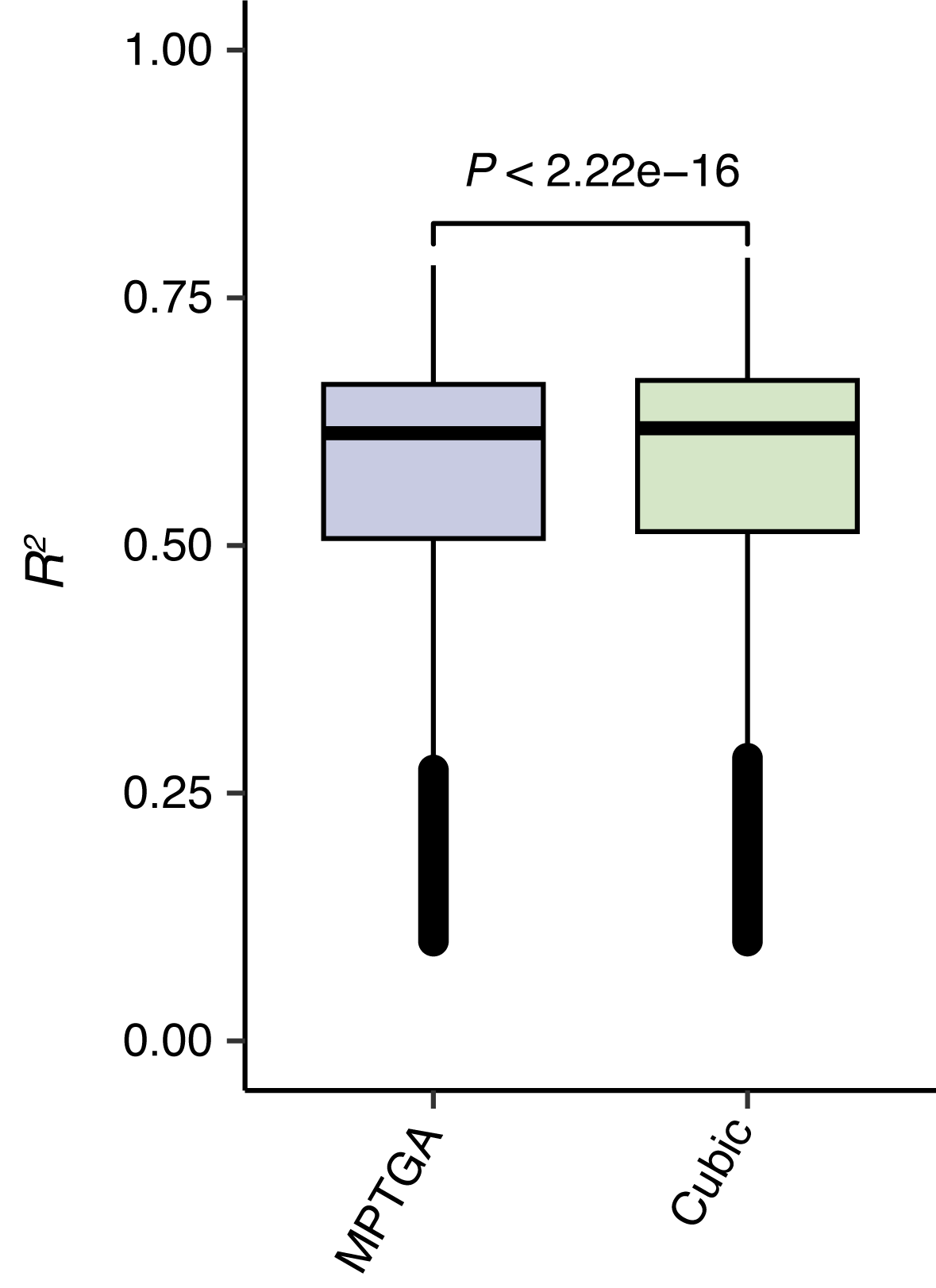

#### Figure S16. The pipeline for ROSMAP cohort quality control and genotype imputation.

Quality control was conducted using PLINK; genotype imputation was performed using Eagle2 and Minimac4 on the TOPMed Imputation Server.

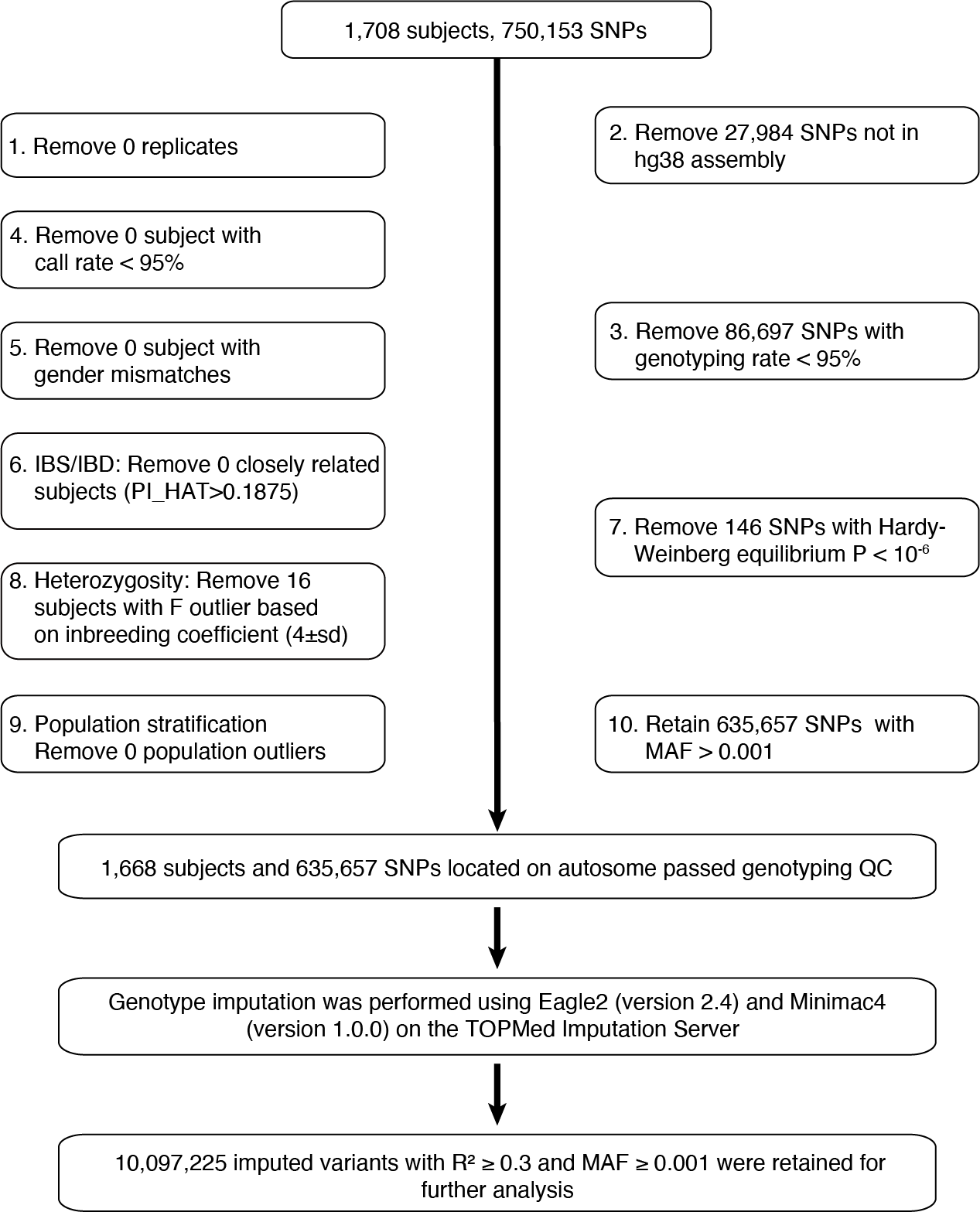

### Supplemental Tables

#### Table S1. Eight significant genotype PCs were leveraged to account for population stratification in the discovery dataset.

| #N | eigenvalue | difference | twstat | *P* value | effect. n |
| --- | --- | --- | --- | --- | --- |
| 1 | 4.78112 | NA | 7.36 | 5.01E-08 | 69.505 |
| 2 | 2.20897 | -2.57215 | 5.354 | 9.76E-06 | 369.698 |
| 3 | 2.11275 | -0.09622 | 11.754 | 1.64E-13 | 632.454 |
| 4 | 1.31151 | -0.80124 | 7.716 | 1.81E-08 | 13771.11 |
| 5 | 1.2357 | -0.07581 | 2.809 | 0.00245681 | 29702.387 |
| 6 | 1.22295 | -0.01275 | 3.735 | 0.000387659 | 41023.73 |
| 7 | 1.1993 | -0.02365 | 1.652 | 0.0183639 | 64090.581 |
| 8 | 1.18925 | -0.01005 | 1.505 | 0.0230912 | 86343.396 |
| 9 | 1.17691 | -0.01234 | -0.195 | 0.206049 | 119055.847 |
| 10 | 1.17183 | -0.00508 | -0.155 | 0.19781 | 144848.088 |
| 11 | 1.16758 | -0.00425 | 0.224 | 0.130827 | 180926.517 |
| 12 | 1.16211 | -0.00547 | NA | NA | NA |
| 13 | 1.15583 | -0.00628 | NA | NA | NA |
| 14 | 1.15558 | -0.00025 | NA | NA | NA |
| 15 | 1.14834 | -0.00724 | NA | NA | NA |
| 16 | 1.14663 | -0.00171 | NA | NA | NA |
| 17 | 1.1433 | -0.00333 | NA | NA | NA |
| 18 | 1.14289 | -0.00041 | NA | NA | NA |
| 19 | 1.13932 | -0.00357 | NA | NA | NA |
| 20 | 1.13848 | -0.00084 | NA | NA | NA |

*P* values is the significance of PCs generated from PLINK*;* *P* values was derived by testing the Tracy–Widom distribution (twstat)^22^ of eigenvalues from the PCA in genotype data; *P* < 0.05 is considered as significance of PC.

#### Table S2. Eight significant genotype PCs were leveraged to account for population stratification in the replication dataset.

| #N | eigenvalue | difference | twstat | *P* value | effect. n |
| --- | --- | --- | --- | --- | --- |
| 1 | 3.4457 | NA | 10.037 | 4.93E-11 | 165.365 |
| 2 | 1.79001 | -1.65569 | 8.501 | 5.50E-09 | 1332.383 |
| 3 | 1.56005 | -0.22996 | 8.335 | 8.95E-09 | 2983.95 |
| 4 | 1.41203 | -0.14802 | 7.502 | 3.38E-08 | 6772.533 |
| 5 | 1.32839 | -0.08364 | 7.019 | 1.29E-07 | 14557.544 |
| 6 | 1.25934 | -0.06905 | 3.202 | 0.00114872 | 31880.685 |
| 7 | 1.23794 | -0.0214 | 2.352 | 0.00566545 | 47386.521 |
| 8 | 1.22115 | -0.01679 | 1.229 | 0.0349905 | 68028.261 |
| 9 | 1.20998 | -0.01117 | 0.509 | 0.0929717 | 91407.364 |
| 10 | 1.19799 | -0.01199 | -1.571 | 0.596377 | 118558.708 |
| 11 | 1.19457 | -0.00342 | -1.348 | 0.525269 | 130621.293 |
| 12 | 1.19249 | -0.00208 | NA | NA | NA |
| 13 | 1.18601 | -0.00648 | NA | NA | NA |
| 14 | 1.18358 | -0.00243 | NA | NA | NA |
| 15 | 1.17705 | -0.00653 | NA | NA | NA |
| 16 | 1.17423 | -0.00282 | NA | NA | NA |
| 17 | 1.17075 | -0.00348 | NA | NA | NA |
| 18 | 1.16487 | -0.00588 | NA | NA | NA |
| 19 | 1.16413 | -0.00074 | NA | NA | NA |
| 20 | 1.16151 | -0.00262 | NA | NA | NA |

*P* values is the significance of PCs generated from PLINK*;* *P* values was derived by testing the Tracy–Widom distribution (twstat)^22^ of eigenvalues from the PCA in genotype data; *P* < 0.05 is considered as significance of PC.

#### Table S3. A list of temporal genetic causal association on PD progression related blood gene expressions for CSF protein levels varying over time using 2sGen-GPS.

*P* values were calculated via VAR model in 2sGen-GPS with *F-test*; discovery dataset, PPMI; replication dataset, PDBP; *P*-value threshold of 5.0×10^-5^ is considered as significance.

Abbreviations: Pval_dis, Pval _rep, Pval _comb, *P* value in the discovery, replication and combined datasets; Coef1_dis, Coef1_rep, Coef1_comb, coefficient of the first order lagged gene series term in the discovery, replication and combined datasets.

#### Table S4. A list of temporal genetic causal association on PD progression related blood gene expressions for neuroanatomic features varying over time using 2sGen-GPS.

*P* values were calculated via VAR model in 2sGen-GPS with *F-test* on one dataset, PPMI; *P*-value threshold of 5.0×10^-5^ is considered as significance.

Abbreviations: Pval, *P* value; Coef1, coefficient of the first order lagged gene series term; Neuroanatomic mrGrad, Neuroanatomic microstructural gradients

#### Table S5. A list of the perturbagen compounds on specific CMap cell line which gene signatures connect to the causal genes of cognitive decline.

pert_iname is the perturbagen name, cell_iname is the cell line in CMap, pert_type is the perturbagen type, fdr_q_nlog10 represents log_10_(FDR) that calculated in CMap, norm_cs represents normalized connectivity score computed by CMap. See https://clue.io/ for the other explanation of indexes.

#### Table S6. Comparison of five temporal eQTL mapping approaches using the simulated dataset with varying over auto-correlation.

*P* values were calculated via *DeLong's* test^9^ for two correlated ROC curves.

#### Table S7. Dynamic *cis*-eQTL SNP annotation and variant effect prediction.

The variant annotation and effect prediction were directly generated from *snpEff* using human genome version of *GRCh38.86.*

#### Table S8. Additive *cis*-eQTL SNP annotation and variant effect prediction.

The variant annotation and effect prediction were directly generated from *snpEff* using human genome version of *GRCh38.86.*

See additional files for Table S3-S8.

### Supplemental Data

#### Data 1. Blood temporal *cis*-eQTLs at FDR<0.05 comprising nonlinear dynamic and additive *cis*-eQTLs .

To obtain the list of nonlinear dynamic *cis*-eQTL, select the rows with ‘Interaction_FDR’ < 0.05 and ‘Interaction_Pval_rep’ < 0.05; to obtain the list of additive *cis*-eQTL, select the rows with ‘Interaction_FDR’ > 0.05 and ‘Interaction_Pval_rep’ > 0.05; position based on human genome version of GRCh38; *P* values from temporal eQTL mapping model with likelihood ratio test; Interaction *P* values from dynamic eQTL mapping model with likelihood ratio test; discovery dataset, PPMI; replication dataset, PDBP.

Abbreviations: Chr., chromosome; FDR, false discovery rate from multiple testing based on Benjamini and Hochberg adjustment method; r_dis, r_rep, auto-correlation proportion in the discovery and replication datasets; Pval, *P* value；PVE, proportion of variance in phenotype explained by temporal eQTL variant; b0k, b1k, b2k (k = 0,1,2,3), coefficient of cubic polynomial in genotype =0,1,2.

#### Data 2. Prioritized genes from the temporal genetic causality analysis using 2sGen-GPS on blood temporal *cis*-eQTLs for neurodegenerative-related phenotypes.

*P* values from VAR model in 2sGen-GPS with *F-test*; discovery dataset, PPMI for PD progression and MAP for cognitive decline; replication dataset, PDBP and SURE-PD3 for PD progression and ADNI for cognitive decline; Importance from integrating all causal genes for a phenotype outcome with permutation-based method.

Abbreviations: Pval_dis, Pval _rep, Pval_comb, *P* value in the discovery, replication and combined datasets; Pval_adjust, Pval_dis adjusted by multiple testing based on Bonferroni adjustment method; Coef*_dis, Coef*_rep, Coef*_comb, lagged correlation coefficient of the * order lagged gene series term in the discovery, replication and combined datasets; SC_rep, Schwarz criterion in the replication dataset.
